# Electrode positioning errors reduce current dose for focal tDCS set-ups: Evidence from individualized electric field mapping

**DOI:** 10.1101/2024.02.19.24302917

**Authors:** Filip Niemann, Steffen Riemann, Ann-Kathrin Hubert, Daria Antonenko, Axel Thielscher, Andrew K. Martin, Nina Unger, Agnes Flöel, Marcus Meinzer

## Abstract

**Objective:** Electrode positioning errors contribute to variability of transcranial direct current stimulation (tDCS) effects. We investigated the impact of electrode positioning errors on current flow for tDCS set-ups with different focality.

**Methods:** Deviations from planned electrode positions were determined using data acquired in an experimental study (N=240 datasets) that administered conventional and focal tDCS during magnetic resonance imaging (MRI). Comparison of individualized electric field modeling for planned and empirically derived “actual” electrode positions was conducted to quantify the impact of positioning errors on the electric field dose in target regions for tDCS.

**Results:** Planned electrode positions resulted in higher current dose in the target regions for focal compared to conventional montages (7-12%). Deviations from planned positions significantly reduced current flow in the target regions, selectively for focal set-ups (26-30%). Dose reductions were significantly larger for focal compared to conventional set-ups (29-43%).

**Conclusions:** Precise positioning is crucial when using focal tDCS set-ups to avoid significant reductions of current dose in the intended target regions.

**Significance:** Our results highlight the urgent need to routinely implement methods for improving electrode positioning, minimization of electrode drift, verification of electrode positions before and/or after tDCS and also to consider positioning errors when investigating dose-response relationships, especially for focal set-ups.

## 1. Introduction

Research on transcranial direct current stimulation (tDCS), a widely used non-invasive brain stimulation technique, has revealed substantial variability of behavioral and neural stimulation effects within and between participants and studies (Ziemann and Siebner, 2015). The underlying sources of this variability are thought to be multifactorial and can broadly be classified as participant and brain stimulation dependent (Fertonani and Miniussi, 2017). The former include trait- and state-dependent characteristics of the participants (Aberra et al., 2018; Antonenko et al., 2019a; Hordacre et al., 2017), that are difficult to account for in data acquisition or analysis. Several stimulation dependent factors (e.g., tDCS timing or duration) are determined by the experimental set-up. However, accurate positioning of electrodes on the participants’ head, one of the most critical stimulation dependent factors, can be affected by experimenter error (i.e., electrode misplacement) or electrode drift over the course of the experiment.

For example, a recent computational modeling study (Woods et al., 2015) demonstrated that 5% drift of large conventional rubber electrodes from their initial positions, equaling 1-1.5 cm on an average head, significantly altered the distribution of the electric field. Comparison of current flow simulations for “actual” electrode positions (derived from magnetic resonance imaging, MRI) with “virtual” electrodes representing “planned” montages, revealed less pronounced current flow in the latter (Indahlastari et al., 2023). Therefore, it was suggested that computational models of tDCS-induced current flow could be improved by accurately representing actual electrode positions (Opitz et al., 2018).

Electrode positions can be verified in studies administering tDCS during MRI (Gbadeyan et al., 2016b; Meinzer et al., 2012; Ulm et al., 2015). However, the vast majority of previous tDCS studies did not administer the stimulation during fMRI (Ekhtiari et al., 2022; Ghobadi-Azbari et al., 2021) and the location of electrodes relative to intended target brain regions remained unknown. With few exceptions (Antonenko et al., 2019b; Indahlastari et al., 2023; Woods et al., 2015), actual electrode positions have also not been considered in computational studies (Hunold et al., 2023). Therefore, electrode positioning errors may explain some of the mixed results in experimental tDCS studies and computational studies investigating associations between current dose and behavioral, neurophysiological and neurochemical outcomes (Hunold et al., 2023). Moreover, errors in electrode placement may be particularly detrimental when using focal set-ups that constrain the current flow to circumscribed brain regions (Gbadeyan et al., 2016a; Martin et al., 2020; Villamar et al., 2013). However, the impact of deviations of electrodes from planned positions on current flow has not yet been examined for focal set-ups, nor have potential differential effects on focal and conventional set-ups been quantified.

This was addressed in the present study that aimed to (a) determine the degree of electrode positioning errors under routine, yet highly standardized, experimental conditions and, (b) quantify potential effects on the electric field dose reaching brain target regions for tDCS set-ups with different focality. We analyzed data acquired in a large concurrent tDCS-fMRI study that investigated behavioral and neural effects of conventional and focal tDCS set-ups targeting two different brain regions. Structural imaging data was used to determine the degree of electrode displacement from planned positions. Individualized current flow modeling was conducted for actual and planned electrode positions, to quantify the effect of positioning errors on the current dose reaching the target regions for both conventional and focal montages.

## 2. Materials and methods

### 2.1. Participants and procedures

Data of 120 healthy young adults (63 men, 57 women, mean±SD age: 22.42±2.58 years) were included. All participated in a study that investigated behavioral and neural correlates of tDCS that was administered during fMRI using two different montages (conventional, focal) and target areas (left inferior frontal, IFG; left primary motor cortex, M1). Thirty participants were scanned with each montage and target area. Groups were comparable regarding sex and age (conventional IFG: 15 men, age mean±SD 23.00±2.92; focal IFG: 17 men, 22.70±2.34; conventional M1: 14 men, 22.2±2.70; focal M1: 14 men, 21.63±2.33; Χ^2^=0.8, p=.85; F(3,116)=1.61, p=.0.19). The study was approved by the local ethics committee and conducted in accordance with the Helsinki declaration. Written informed consent was obtained prior to study inclusion.

Stimulation was administered in double-blinded, balanced cross-over designs with two sessions, during which either anodal tDCS (1 mA) or sham stimulation (0.05 mA, sinusoidal) was applied for 24 min during resting-state and task-based fMRI (Note: FMRI data will be reported elsewhere). Afterwards, T1-weighted images were acquired with electrodes attached, to allow verification of actual electrode positions. Additional T1- and T2-weighted images without electrodes were acquired for individualized electric field modeling based on planned and empirically determined actual electrode positions. Procedures were identical for active and sham tDCS and staff administering tDCS were blinded (using pre-assigned codes triggering active or sham tDCS). Hence, MRI and simulation data from both sessions were used (N=240 datasets).

TDCS was administered using an MRI-compatible multi-channel stimulator (DC-Stimulator MC-MR, NeuroConn). Planned electrode positions are illustrated in **Figure 1A-D**). The 10-20 EEG system and an EEG cap were used for targeting. M1 corresponded to the C3 position (Gbadeyan et al., 2016b), IFG was determined as described previously (Meinzer et al., 2012). For ***conventional montages,*** rectangular 5×7 cm^2^ rubber electrodes were placed in saline-soaked sponge pockets. Anodes were centered over the respective target areas, the longer edges horizontally aligned with the 20% isodistance line (Arena et al., 2021). Cathodes were placed in 10×10 cm^2^ sponge pockets, positioned over the right supraorbital cortex (FP2) and vertically aligned along FP2-F2. Electrodes were held in place by rubber straps. ***Focal montages*** used circular rubber electrodes (radius=1.25 cm) affixed to the scalp with ∼1 mm of Ten20 conductive paste and held in place with EEG caps. A printed 3D template assured consistent application of conductive paste (**Figure 1E**). Anodes were centered over the same position as the conventional electrodes and were surrounded by three equally spaced cathodes. Positioning of the cathodes was standardized with another 3D printed template with a radius of 4.5 cm (anode-cathode centers, **Figure 1F**). After determining the position of the anode, one of the cathodes was placed along the vertically aligned connection of T7-LPA (IFG) or C3-C1 (M1), **Figure 1C-D**.

**Figure 1.**
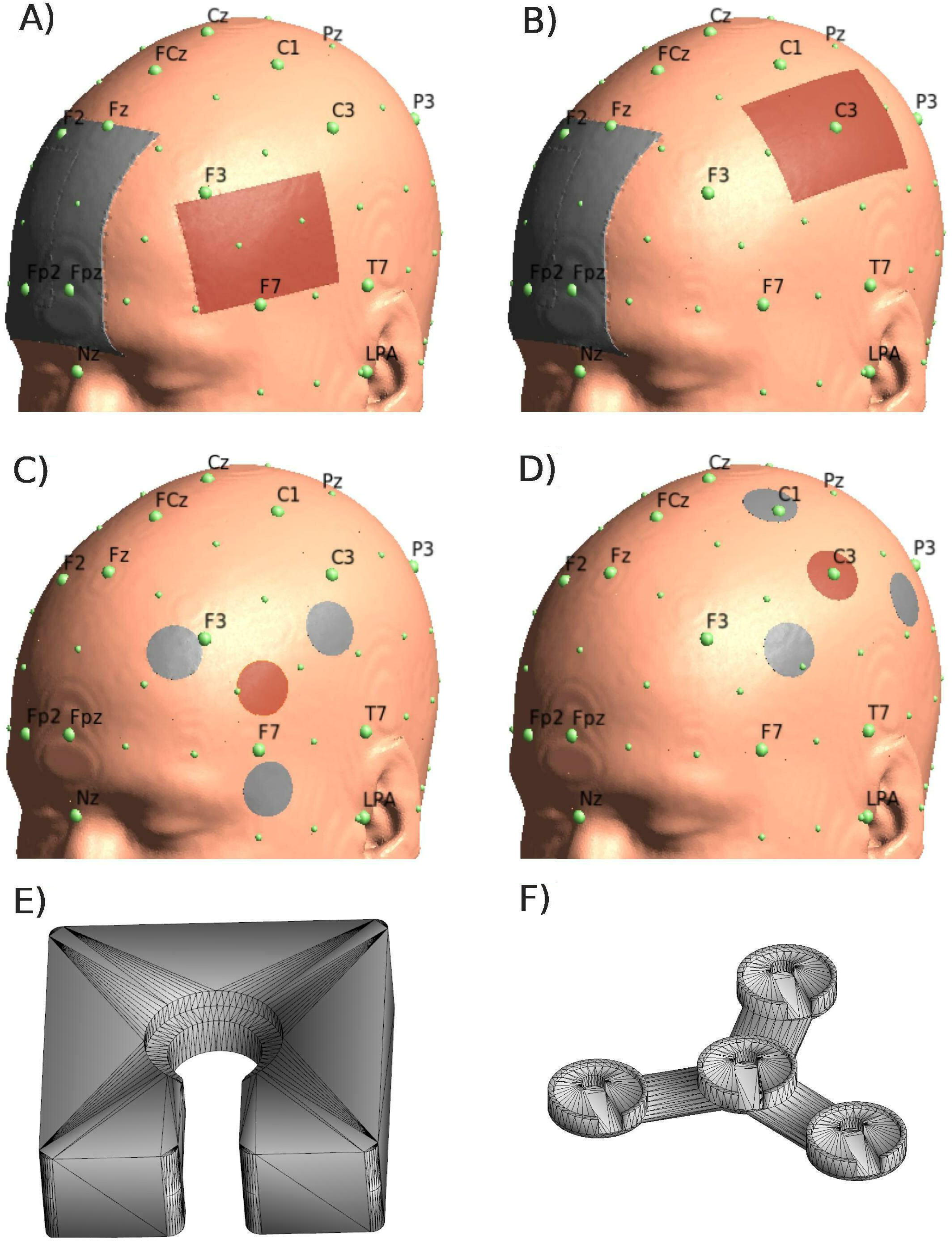
Planned transcranial direct current (tDCS) montages. Illustrates the planned conventional and focal montages (cathodes=blue, anodes=red) and relevant 10-20 EEG system coordinates used for localization of the target regions and consistent orientation of electrodes. **Upper panels:** Conventional tDCS montages targeting the A) inferior frontal gyrus (IFG) or B) motor cortex (M1). **Middle panels:** Focal 3×1 tDCS montages targeting the C) IFG or D) M1. **Bottom panels:** Show the 3D models that were used to print E) the device that was used for consistent application of conductive gel, and F) the spacer that was used to ensure consistent placement of cathodes relative to the anodes in the focal set-ups (center-to-center electrode distance: 4.5 cm).

### 2.2. Structural MRI

MRI data were acquired with a 3-Tesla Siemens Verio scanner at the University Medicine Greifswald using a 32-channel head coil. After functional imaging, T1-weighted images with the electrode montages attached to the scalp were acquired to determine electrode positions. Additional T1- and T2-weighted images without electrodes were acquired for current modeling (T1: 1 mm^3^ isotropic voxels, TR=1690 ms, TE=2.52 ms, TI=900 ms, flip angle: 9°; T2: 1 mm^3^ isotropic voxels, TR=12770 ms, TE=86 ms, flip angle: 111°).

### 2.3. Planned and actual electrode positions

To compare the location and the corresponding simulated electric fields between planned (as intended) and empirically derived actual montages (identified using the individual participant’s MRI data), planned coordinates derived from MNI space were transformed and processed in subject space for each participant. Hence, deviations of actual from planned electrode positions and electrical field simulations based on actual and planned positions were both calculated in subject space.

For the planned IFG montages (conventional, focal), the anodes were centered on the MNI52 template at scalp coordinates [x/y/z −69/35/17], corresponding to 10-20 EEG positions described previously (Meinzer et al., 2012). The center positions for M1 montages were equivalent to C3. The longer edges of the 5×7 cm^2^ anodes were horizontally aligned with the 20% isodistance line (Arena et al., 2021). Cathodes for both conventional montages were centered over the right supraorbital cortex (FP2) and vertically aligned along the connection of FP2-F2. Positions of all conventional electrodes were then non-linearly transformed into subject space of each participant via a SimNIBS (Thielscher et al., 2015) internal function (mni2subject_coords). For the focal montages, MNI coordinates of the anodes were transformed into subject space and cathode coordinates were generated using a customized SimNIBS internal function (tDCS_Nx1.py). This function automatically generated equally spaced 3×1 electrode configurations, with a center-to-center radius of 4.5 cm between the anode and each cathode. For both target areas, the center of the first cathode was positioned on a line between the anode center and an orientation point (IFG: −71/34/-10; M1: - 38/-11/87), which aligns the line parallel along the connection of T7-LPA or C3-C1, respectively (**Figure 1A-D**). This corresponds to the planned positioning of the cathodes in the experimental set-up. Coordinates and orientations are detailed in **Supplementary Table A.1**.

### 2.4. Actual electrode positions

T1/T2-weighted images acquired without electrode montages were reoriented to match the orientation of the MNI standard template using fslreorient2std. Afterwards, the T1-weighted image with the actual montage was co-registered to the reoriented T1-image and x/y/z coordinates of the respective electrode centers were manually determined by two independent raters (FN, MM) using the render function of MRIcron (https://www.nitrc.org/projects/mricron). In addition, the orientation of the electrodes along the y-axis was also extracted for the conventional montages using the plug of the connecting cable as reference. Consensus between the two raters was reached if electrode positions differed.

### 2.5. Simulation of electric fields

SimNIBS 3.2 (Thielscher et al., 2015) was used to calculate the electric field induced by tDCS, based on the finite element method and individualized tetrahedral head meshes generated from structural T1- and T2-weighted images (without electrodes) of the participant (Saturnino et al., 2019; Thielscher et al., 2015; Windhoff et al., 2013). SimNIBS 3.2, integrated tools from SPM12, CAT12 (Gaser et al., 2022), FreeSurfer (https://surfer.nmr.mgh.harvard.edu), FMRIB’s FSL (https://fsl.fmrib.ox.ac.uk/fsl/fslwiki), MeshFix (Attene, 2010), and Gmsh (Geuzaine and Remacle, 2009) generated automatic segmentations and head meshes. Head reconstruction for each subject and session was performed using the incorporated headreco tool (Nielsen et al., 2018). Freeview (FreeSurfer) was used for inspection the reconstructed tissue compartments. All datasets were deemed appropriate.

Conventional anodes were defined as rectangle (5×7 cm^2^, 0.2 cm thickness) and a small, dorsally orientated plug (0.5×2 cm^2^) with standard thickness values from SimNIBS, inserted into sponge pockets (5×7 cm^2^, 0.8 cm thickness). Cathodes were centered in the middle of a 10×10 cm^2^ sponge pocket (thickness 0.8 cm). The simulated current was set to 1 mA for the anode, −1 mA for the cathode. Rubber electrodes of the focal montages were defined as an ellipse (1.25 cm radius, 0.2 cm thickness). The applied current 1 mA for the anode, the sum of the currents for the three cathodes was −1 mA. Standard conductive values from SimNIBS were used. Afterwards, surface overlays of the middle layer of the cortical sheet were estimated from the pial and white matter surfaces and transformed into “fsaverage” space (Freesurfer). For planned and actual coordinates, the vector norm of the electric field |E| (i.e., magnitude E) and the component of the electric field oriented normally to the cortical sheet nE (Antonenko et al., 2019b) were extracted from each transformed middle layer. Note: All field simulations were conducted in subject space. However, for illustrative purposes (**Figure 3**; **Supplementary Figure A.5**), weighted group averages and standard deviations of the electric field strength and the normal component were calculated for each montage in “fsaverage” space.

### 2.6. Region-of-interest (ROI) analysis

To investigate if planned and actual electrode coordinates differed with regard to the respective simulated electrical fields in the target regions, data was extracted from spheres with a radius of 1.25 cm (i.e., radius of focal-tDCS electrodes) or 2.5 cm (2x radius) overlapping with the cortical surface of the two target regions for each montage. Two sphere sizes were used to investigate the regional precision of stimulation effects (Mikkonen et al., 2020).

The M1 ROI was centered at MNI coordinates −52/-16/58, i.e., the cortical projection point of C3 (Okamoto et al., 2004). Accuracy of the gray matter surface (x) coordinate was controlled for with MRICron. Please note, the location of the electrode center targeting the left IFG did not correspond to a standard 10-20 EEG system position, but was based on a previous study of our group (Meinzer et al., 2012). To identify the optimal projection point, we used the SimNIBS electrode creation function to simulate small line-shaped electrodes on the head to reconstruct relevant 10-20 EEG positions described in this previous study. This allowed us to determine the planned target scalp coordinates. We then used the SimNIBS „grey matter to head“ projection tool to identify the optimal grey matter projection point (−57/24/12), corresponding to the closest projection towards the center of the planned target scalp coordinates.

The average E-field magnitude was extracted from the cortical surface overlapping with the respective ROIs. For the normal component, only positive values were used (Antonenko et al., 2019b). For illustrative purposes (i.e., to visually present the variability of actual electrode positions across all participants), MNI center coordinates were also transformed into fsaverage space (https://surfer.nmr.mgh.harvard.edu/fswiki/CoordinateSystems) via affine transformation.

### 2.7. Statistical analysis

Linear mixed models (LMMs, (Verbeke and Molenberghs, 2000)) were calculated to evaluate deviations of actual from planned electrode positions and differences between the (simulated) induced electric fields for actual and planned montages. For LMM parameters see **Supplementary Tables A.2, A.4 and A.6**. To find the best model, all parameters of interest were initially added as interaction of fixed effects and subjects as random effects, with parameters as additional random slope within subjects (e.g., to account for individual differences of electric field magnitude in actual vs. planned electrode positions). Subsequently, parameters or interactions of less importance were removed and models were compared using likelihood ratio tests and the Bayesian information criterion (BIC). The model with the smallest BIC was selected, yielding a sufficient number of parameters while avoiding overfit.

The R-package (R Core Team, 2022) was used for statistical analyses; including the lme4 package (Bates et al., 2015) for computing p-values based on conditional F-tests with Kenward-Roger approximation. Effect sizes (semi partial R²) were computed for fixed effects in the LMMs (Jaeger, 2017) using the r2glmm package with the Nakagawa-Schielzeth approach, using fixed and random effects for determining explained variance (Nakagawa and Schielzeth, 2013). Note that in an LMM only simple effects and simple contrasts are provided, meaning the effect of one factor is dependent on the level of the reference model of all other given factors. Then, the estimated marginal means (EMMs) were calculated for main effects, i.e., the internal comparison of one factor level over all other factor levels. The interaction of significant main effects with other factor levels was determined using simple effects and contrasts. To estimate main effects, the emmeans package (Lenth et al., 2023) was used. EMMs and post-hoc tests were calculated, applying the Tukey correction for multiple comparisons.

### 2.8. Electrode position differences

To test for deviations between planned and actual electrode positions, differences between the respective coordinates (in mm) were calculated for each spatial dimension (x/y/z coordinates) in subject space. For illustrative purposes, actual coordinate positions were transformed from subject-to-MNI space. Absolute differences between planned and actual positions of the center anodes were used as dependent variable for the LMM; montages (conventional, focal), area (IFG, M1) and spatial dimension (x/y/z), were entered as fixed effects [N=30 participants/group x 2 sessions x 2 areas x 2 montages x 3 spatial dimensions]. Random intercepts were fitted for each subject. Models were adjusted for sex, session (day_1,2_) and stimulation type (active/sham). Interaction effects were compared using EMMs and post-hoc paired t-tests.

### 2.9. Magnitude E

In order to investigate the impact of individual deviations in electrode positioning on the simulated induced current, (a) weighted mean electric field values were calculated using SimNIBS for planned and actual electrode positions and (b) extracted from spherical ROIs (1.25 and 2.5 cm radius) overlapping with cortical volumes in the left IFG and M1 as described above (**Figure 3E-H**). Stimulation site, montages and ROI radius were entered as fixed effects in the LMM, participants as random effects with variation of the slope for ROI radius and electrode position, [N=30 participants x 2 sessions x 2 areas x 2 montages x 2 ROI radius x 2 electrode position]. Subjects were random effects, ROI radii and electrode positions random slopes within subjects. Interaction effects were compared with EMMs and post-hoc paired t-tests. Data analysis for the normal component of the electrical field was identical. Results were comparable to the magnitude E analysis and only the most relevant outcomes are reported (see **Supplementary Materials** for details).

## 3. Results

### 3.1 Electrode positions

The actual positions of the anode centers for individual participants and all montages are illustrated in **Figure 2A-D**. Detailed information on x/y/z coordinates and deviations of actual from planned positions for all electrode centers are provided in **Supplementary Figures A1-4**. For both IFG montages, mean displacement was >1 cm in ventral and posterior directions (**Supplementary Figures A1-2**). Mean displacement for M1 montages was observed mainly in the anterior direction (on average <1 cm, **Supplementary Figures A3-4**). Thus, systematically better placement accuracy was achieved for the easier to localize M1 montage (**Figure 2E**; 4.47 mm, 95%-CI: [3.55, 5.39]; t=11.71, p<0.001). No significant differences were found for the main effect of montage ( **Figure 2F**). Results of the linear mixed model analysis for the dependent variable electrode position difference (planned-actual) are detailed in **Supplementary Table A.2**, interactions of the main effects in **Supplementary Table A.3**.

**Figure 2.**
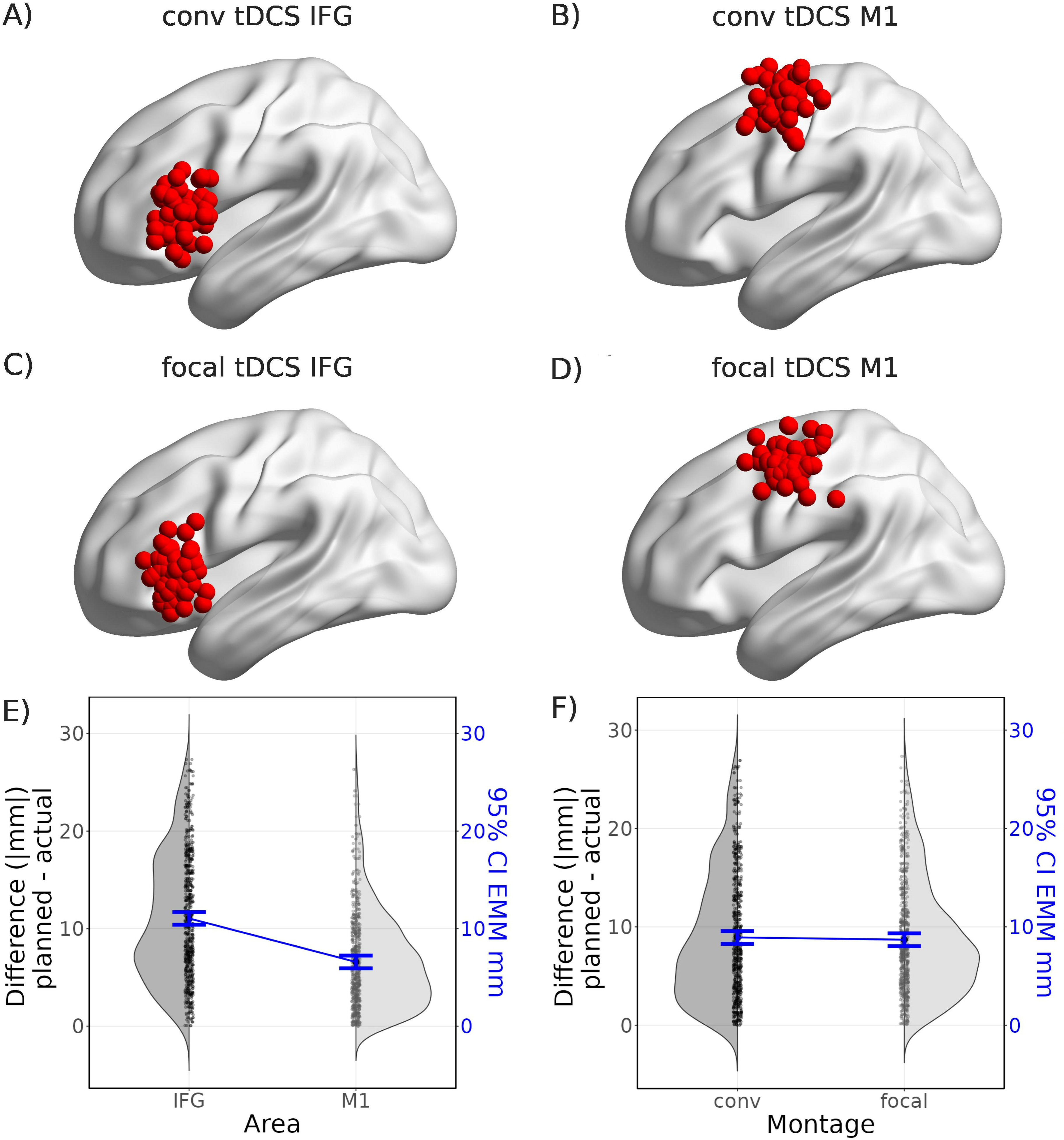
Empirically determined positions of anodes for all montages. Red spheres illustrate the empirically determined actual positions of the center anodes for all participants and sessions (Note: for illustrative purposes, data was transformed into standard space). **Upper panels:** Conventional tDCS montages targeting the A) inferior frontal gyrus (IFG) or B) motor cortex (M1). **Lower panels: Middle panels:** Focal tDCS montages targeting the C) IFG or D) M1. **Bottom panels:** Illustrate differences between planned and actual electrode positions in |mm| for the factors E) area and F) montage for individual participants and distribution as half violin plots (grey); scale on the left y-axis. In blue, estimated marginal means (EMMs) of the corresponding factors with 95% confidence intervals (CI) in V/m; scale on the right y-axis. For additional details see **Supplementary Table A.3**; conv=conventional.

**Figure 3.**
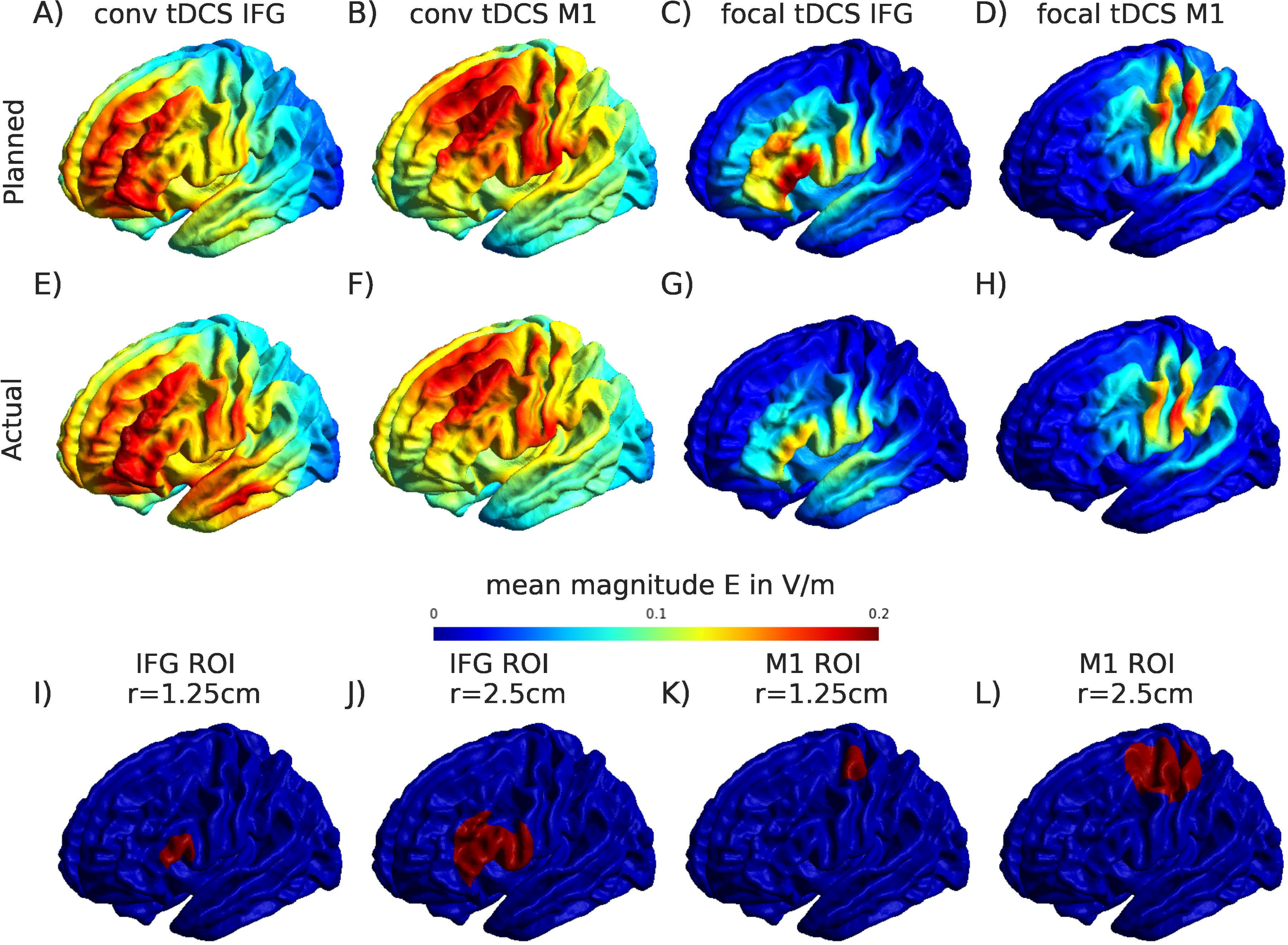
Electric field distribution of magnitude E for actual and planned montages. Average of the electric field distribution across participants for the four montages derived from finite element method calculations using SimNIBS. Magnitude E (|E|) averages (in V/m) are shown (Note: Only the left side of the brain is shown). **Upper panels:** Results for planned coordinates for conventional tDCS montages targeting the A) inferior frontal gyrus (IFG) or B) motor cortex (M1), and focal tDCS montages targeting the C) IFG or D) M1. **Middle panels:** Results for actual electrode positions for conventional tDCS montages targeting the A) IFG or B) M1 and focal tDCS montages targeting the C) IFG or D) M1. **Bottom panel:** Illustrates the overlap between the cortical surface of the target regions and spheres with two different radii (r=1.25/2.5 cm) that were used to extract data for the region-of-interest (ROIs) analyses for the I-J) IFG or K-L) M1, respectively.

### 3.2 Electric field strength

Magnitude E is visualized for planned (**Figure 3A-D**) and actual electrode positions (**Figure 3E-H**) for each montage. As expected, higher field magnitudes across the entire brain were observed for conventional compared to focal montages for planned electrode positions (95^th^ percentile mean±SD V/m conventional/focal: IFG 0.146±0.016/0.069±0.014, t(df=58)=27.36, p<0.001; M1 0.156±0.019/0.061±0.014, t(58)=31.59, p<0.001). A similar pattern was confirmed for actual electrode positions (conventional/focal IFG: 0.146±0.002/0.057±0.012, t(58)=58.56, p<0.001, M1: 0156±0.019/0.051±0.061, t(58)=68.34, p<0.001).

To investigate current dose in the target regions, ROIs with two different sizes (radii: 1.25/2.5 cm) were used to extract magnitude E from the cortex overlapping with the two target areas (**Figure 3I-L**) for planned and actual positions. Results of the LMM for different montages, electrode positions and ROI radii are shown in **Table A.4**. Estimated marginal means (EMMs) of fixed effects from the LMM model were calculated to compare main effects and simple contrasts (**Table A.5**).

There was no significant difference in magnitude E between the target areas IFG and M1 (EMM difference=0.01 V/m, 95%-CI: [-0.10, 0.02]; t(115)=1.12, p=0.263; **Figure 4A**). However, the simulated electric field strength was significantly higher for planned compared to actual electrode positions (0.02 V/m, 95%-CI: [0.01, 0.02]; t(826)=12.56, p<0.001; **Figure 4B**) and also in the smaller ROI (0.018 V/m, 95%-CI: [0.0155, 0.0199]; t(826)=16.96, p<0.001; **Figure 4C**). This effect was most pronounced between montages, showing significantly higher magnitude E for conventional compared to focal montages (0.03 V/m, 95%-CI: [0.02, 0.04]; t(115)=5.68, p<0.001; **Figure 4D**).

**Figure 4.**
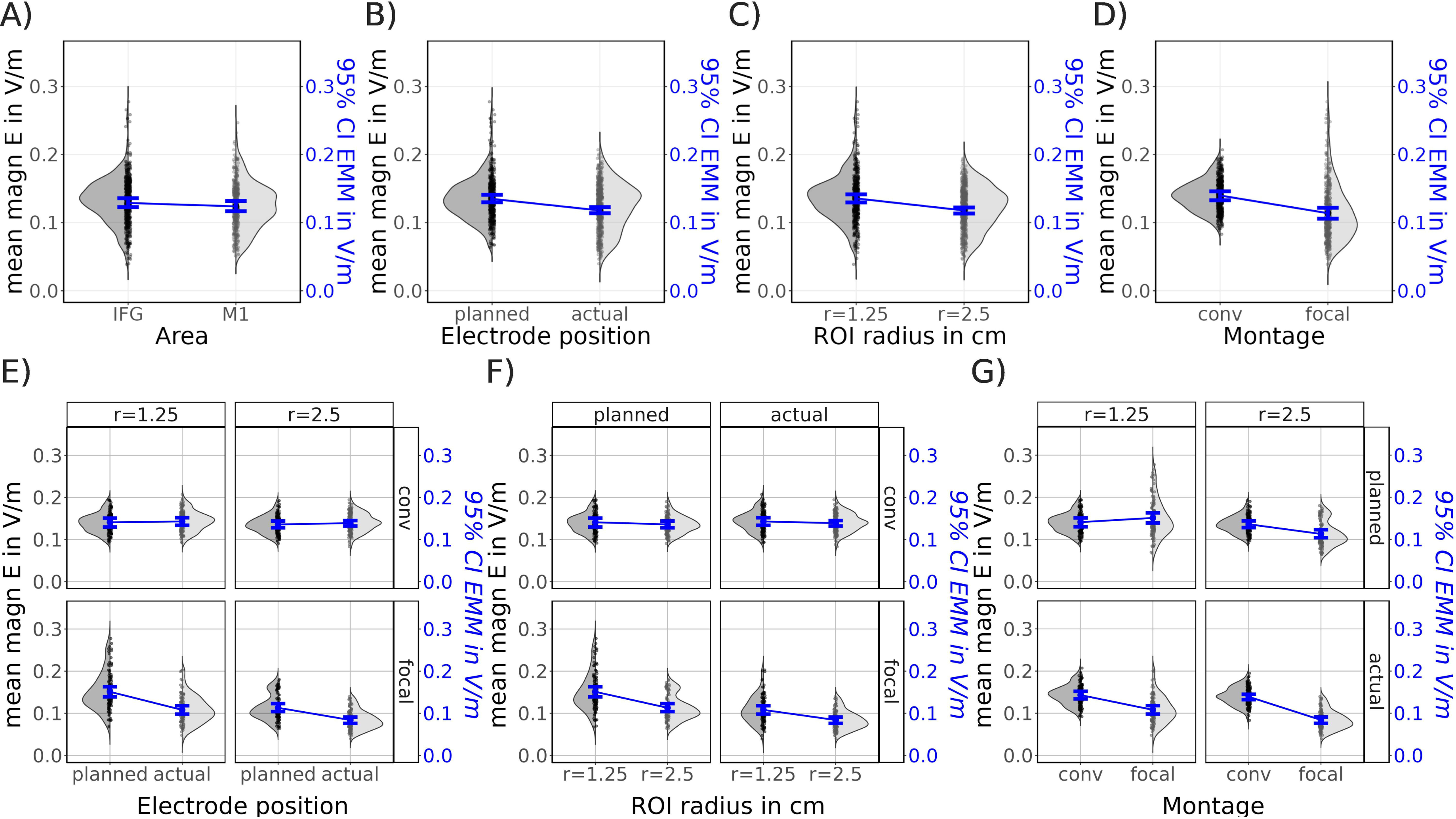
Main effects and simple contrast of the electrical field strength. **Upper panels:** Illustrate the results of the electrical field strength (magnitude E) analysis for the factors A) area, B) electrode position C) region-of-interest (ROI) radius (r) and D) montage. In gray, mean magnitude E values in V/m for individual subjects and distribution as half violin plots; scale on the left y-axis. In blue, 95% Confidence Interval (CI) in V/m of the estimated marginal means (EMMs), scale on the right y-axis. **Bottom panels:** Simple contrasts for factors E) electrode position, F) ROI radius and G) montage with interaction factors (shown in boxes, top and right side). For additional details see **Supplementary Table A.5**; magn=magnitude.

The interaction behind the main effects revealed that electrode positioning errors significantly decreased magnitude E for the focal montages in both ROIs (**Figure 4E** lower panels: r=1.25, 0.04 V/m 95%-CI: [0.04, 0.05]; t=27.99, p<0.001, df=826; r=2.5, 0.03 V/m, 95%-CI: [0.02, 0.04]; t(826)=19.40, p<0.001), resulting in a dose reduction of 26.7% and 27.3%. Positioning errors did not affect magnitude E for the conventional montages (**Figure 4E** upper panels: r=1.25: 0.003 V/m, 95%-CI: [-0.01, 0]; t(826)=-1.99, p=0.127; r=2.5: 0.004 V/m, 95%-CI: [-0.01, 0]; t(826)=-2.50, p=0.056).

Direct comparison of the two different ROI radii allowed further assessment the regional precision of the stimulation. Increasing the ROI radius resulted only in a small reduction of magnitude E for the conventional montages (**Figure 4F** upper panels: planned 0.01 V/m, 95%-CI: [0, 0.01]; t(826)=3,32, p=0.003, df=826; actual 0.004 V/m, 95%-CI: [0, 0.01]; t(826)=2.810, p=0.010). As expected, significant decreases in magnitude E in the larger ROI confirmed more circumscribed current delivery for planned and actual electrode positions in the focal set-ups (**Figure 4F** lower panels; planned: 0.04 V/m, 95%-CI: [0.03, 0.04]; t(826)=24.62, p<0.001, actual: 0.02 V/m, 95%-CI: [0.02, 0.03]; t(826)=16.02, p<0.001).

Notably, with *planned positioning* of the electrodes, focal montages tended to induce a higher magnitude E in the immediate target region (r=1.25 cm) than conventional montages (**Figure 4G**, top left panel: −0.01 V/m, 95%-CI: [-0.028, 0.01]; t(136)=-1.73, p=0.09; 7% increase). The higher precision of the focal compared to conventional set-ups is illustrated by a significant decrease of magnitude E in the larger ROI (**Figure 4G** top right: 0.02 V/m, 95%-CI: [0.01, 0.04]; t(136)=4.81, p<0.001). Most importantly, *deviations from the planned electrode positions* resulted in a significant decrease of magnitude E for focal compared to conventional montages for both ROI radii (**Figure 4G**; bottom left: r=1.25, 0.04 V/m, 95%-CI: [0.02, 0.05]; t(136)=7.22, p<0.001; bottom right: r=2.5, 0.06 V/m, 95%-CI: [0.05, 0.07]; t(136)=15.43, p<0.001), corresponding to a dose reduction of 28.6% and 42.9%.

Results for the normal component of the electric field (nE) largely confirmed the above findings, all effects went in the same direction. Details and statistics are reported in the **Supplementary Materials** and **Supplementary Figures A.5-6**. The most important findings were that (a) planned positions for focal compared to conventional montages resulted in significantly higher nE in the immediate target region (**Figure A.6G**, left upper panel) and, (b) using actual electrode positions resulted in a significant decrease of nE for the focal compared to the conventional montages in both ROIs (**Figure A.6**, lower panels). For additional information regarding the focality of the stimulation, please see **Supplementary Materials** (Section A.3) and **Supplementary Figure A.7**, where we present additional ROI analyses comparing the results of electrical field simulations for the immediate target region (r=1.25) with those of the surrounding areas only (i.e., ROI r=2.5 minus the smaller center ROI).

## 4. Discussion

We analyzed data from a large intrascanner tDCS study, which allowed quantification of electrode displacement from intended positions for conventional and focal montages. Individualized current modeling for planned and actual electrode positions allowed determining the impact on simulated current flow to the respective target regions. There were no significant differences in placement accuracy between conventional and focal montages. However, significantly larger deviations were observed for the more difficult to locate IFG montages. Several key findings emerged from the current modeling analyses: First, optimal positioning of the focal montages increased current flow to the target regions relative to conventional montages (7% |E|, 12.5% nE). Second, deviations from planned positions did not substantially affect current flow to the target regions for the conventional montages. Third, positioning errors significantly reduced the induced current in both ROIs for focal montages (26.7-27.3% |E|, ∼30% nE). Fourth, dose reductions were significantly more pronounced for focal vs. conventional set-ups (28.7-42.9% |E|, 25-42.9% nE). Results were highly consistent for both components of the electrical field. This suggests that precise positioning of electrodes is particularly important when using focal set-ups, to avoid significant reductions in current flow to the intended target regions. Our results also highlight the need for taking into account actual electrode positions in computational tDCS studies, especially for focal montages.

Deviations of electrodes from intended positions may be a central factor underlying variability of tDCS effects in experimental and computational studies (Indahlastari et al., 2023; Woods et al., 2015). Our data confirmed substantial deviations from intended montages under routine, yet highly standardized experimental conditions. Deviations were more pronounced for IFG montages (>1 cm in ventral/posterior directions), thereby exceeding recommendations for negligible placement errors for conventional montages (Indahlastari et al., 2023). Notably, the margin for errors is likely smaller for focal set-ups due to their higher regional precision. This is supported by our modeling results, demonstrating that deviations from intended electrode positions resulted in significantly reduced current flow to both target regions for focal vs. conventional set-ups, even though errors were <1 cm for the M1 montages (Rich and Gillick, 2019).

The results of our computational analyses are largely in line with previous studies, demonstrating that electrode drift (Woods et al., 2015) or placement inaccuracy (Indahlastari et al., 2023) can significantly affect outcomes of computational models. However, these studies only considered conventional montages and quantified changes in current distribution across brain regions (Woods et al., 2015) or the entire brain/head (Indahlastari et al., 2023). We substantially extended these findings by comparing effects of electrode positioning errors between conventional and focal montages and by focusing the analyses of effects specifically on the two target regions. This is of particular interest, because several computational modeling studies have suggested that the electric field dose reaching target regions is a key factor for stimulation outcome (Hunold et al., 2023). Indeed, by using a ROI approach to characterize current dose in the target regions, we demonstrated that focal compared to conventional set-ups can induce a higher electric field dose in the target regions with optimal positioning, but also that the former are disproportionately more affected by electrode positioning errors. The resulting significant drop in electric field dose in the target regions emphasizes the urgent need to assure accurate electrode placement and preventing drift when using focal set-ups. In line with previous studies (e.g., Bai et al., 2014; Bhattacharjee et al., 2019), we also demonstrated that the electric field dose in the target regions for conventional montages is relatively unaffected by electrode positioning errors due to the wider distribution of the induced current. While this lack of focality renders them less useful for revealing regionally specific causal brain-behavior relationships, this may have advantages in contexts where experimenter error is more likely to occur (e.g., routine clinical care, multicenter intervention studies).

## 5. Limitations

Results are restricted to the montages and target regions used; effects may differ for other set-ups. However, we selected common montages and position error effects were largely consistent for the target regions. We cannot quantify how representative the degree of positioning errors in our study are. Indeed, such errors have largely been neglected in experimental and computational tDCS studies and positioning accuracy has rarely been investigated (Antonenko et al., 2019b; Indahlastari et al., 2023; Seibt et al., 2015). Similar to our own study, the majority of previous tDCS studies employed 10-20 EEG scalp coordinates for targeting (Thair et al., 2017), an approach with limited anatomical precision (De Witte et al., 2018). This likely explains the relatively large variability in placement accuracy, despite highly standardized procedures. Another contributing factor could be the complex intrascanner procedure, which requires participants to walk to and be positioned inside the MRI scanner with electrodes attached. Structural imaging was not performed before and after functional imaging, so we cannot determine when the positioning errors occurred. However, irrespective of their origin, our results highlight the impact of this factor on current flow to the target regions, specifically for focal montages. Finally, the target regions for our ROI analyses were motivated by previous tDCS-fMRI studies that demonstrated beneficial stimulation effects on word-retrieval (Meinzer et al., 2012; Meinzer et al., 2023; Darkow et al., 2017; Martin et al., 2017; Meinzer et al., 2014). This allowed us to quantify effects of electrode placement errors on simulated current flow to the intended target regions. However, future analyses of behavioral and functional imaging data of this study are required to determine the functional relevance of current flow reduction to these ROIs.

## 6. Conclusions

Precise positioning of focal montages can induce a higher current dose in target regions compared to conventional montages. However, deviations from planned positions disproportionately affect current dose for focal set-ups. Future studies are advised to routinely implement appropriate methods for improving electrode positioning (e.g., electrode placement guided by neuronavigation; (Jog et al., 2021)), minimization of drift (Woods et al., 2015), verification of electrode positions before and/or after tDCS (Indahlastari et al., 2023; Knotkova et al., 2019), consideration of positioning errors in computational studies investigating dose-response relationships (Hunold et al., 2023).

## Acknowledgements

This research was funded by the German Research Foundation (project grants: ME 3161/3-1; FL 379/26-1; INST 292/155-1 FUGG; Research Unit 5429: FL 379/34-1, FL 379/35-1, ME 3161/5-1, ME 3161/6-1, AN 1103/5-1, TH 1330/6-1, TH 1330/7-1). AT was supported by the Lundbeck foundation (grant R313-2019-622).

## Data statement

The raw MRI data used for efield calculations are not publicly available due to potential identifying information that could compromise participant privacy. Source data are provided in the manuscript and all data used for statistical analyses that are represented in the respective tables and figures can be found here: https://github.com/LawsOfForm/VerFlu_Simulation_Niemann_2023

All analyses were performed using publicly available toolboxes: <u>Anaconda 3.2.31</u>, <u>Python 3.7.13</u>, <u>R</u> <u>version 4.3.1</u>, <u>MATLAB R2021b</u>, <u>SPM12</u>, <u>CONN 20.b</u>, <u>Cat</u>, <u>BrainNetViewer 1.7</u>, <u>SimNIBS 3.2.6</u>, <u>FreeSurfer Version 7.4.1</u>, <u>FSL 6.0.0</u> and <u>gmsh</u>. Customized codes and instructions are available within the respository.

## Abbreviations

E: electrical field
IFG: inferior frontal gyrus
magnitude E: magnitude of electrical field
M1: motor cortex
nE: normal component of the electrical field
SimNIBS: simulation of non-invasive brain stimulation
tDCS: transcranial direct current stimulation
ref: reference model

