## Appendix for "Electrode positioning errors reduce current dose for focal tDCS set-ups: Evidence from individualized electric field mapping"

#### A.1 Electrode differences

For the electrode positioning error analysis (i.e., the comparison of planned vs. actual locations) reported in the main part of the manuscript, only deviations of the anodes were used. For an overview of deviations from planned positioning of the cathodes for all four montages see **Figures A.1-4**. Planned and electrode positions were overlaid on an MNI brain and are illustrated in **Figures A.1-4A** and **C**, respectively. Actual and planned positions for all spatial dimensions and montages are shown in **Figures A.1-4B**. Differences between planned and actual positions in mm are shown in **Figures A.1-4D**. Note, for the linear mixed model (detailed in **Table A.2**), absolute differences between planned and actual positions were calculated and used as dependent variable.

*A.1.1. LMM for electrode differences*

Full results of the linear mixed model analysis for the dependent variable of the difference between planned and actual coordinate position are shown in **Table A.2**. All effects shown in the model are simple effects or simple contrasts, meaning that they have to be interpreted relative to a reference model in which all levels of all factors are set to zero. A reference model provides a baseline for evaluating the significance and contribution of fixed effects or covariates to the model's performance. The reference model for the analysis of deviations between planned and actual electrode positions with the factors area, montage and spatial dimension was: IFG, conventional montage, y dimension. Results showed that the actual position of the conventional IFG electrode was shifted 14.92 |mm| $R^2$=.296 on the y-axis (anterior-posterior axis) regardless of the direction. Changing the level of a factor gives the simple effect of this factor according to the dependent fixed level of all other factors of the reference model. For example, if the IFG conventional y coordinate is contrasted with the M1 conventional y coordinate, the positioning error is reduced by -5.24 mm (95%-CI: [-6.67, -3.81]) (see **Table A.2**). Complex models with many interactions like this one are not easy to interpret, therefore estimated marginal means of the factors can help to understand the model by showing effects between the level of factors, regardless of other parameters in the model. To facilitate the interpretation of the many significant fixed effects and interactions, post-hoc tests of main effects and simple contrasts were calculated and are shown in **Table A.3**.

*A.1.2. EMMs of LMM for electrode differences*

Main effects for area (**Figure 2E)** and montage (**Figure 2F)** are described in the main manuscript. Regarding coordinate dimension, positioning errors were similar in the y and z dimension, showing only small but significant differences between y and z dimensions of 0.93 mm (CI-95%: [0.15, 1.72], t=3.13, p=0.01, df=1192) **Table A.3**. In contrast, there was a significant increase of error rates of nearly 4 mm between x and y or x and z dimension (contrast y-x: 4.49 mm 95%-CI: [3.70, 5.28]; t=15.05, p<0.01, df=1192; contrast z-x: 3.56 mm 95%-CI: [2.77, 4.34]; t=11.92, p<0.001, df=1192; **Table A.3**). Errors in the x direction (later-medial axis) were not considered further, since the x axis is not of major importance in SimNIBS simulations because it is automatically projected onto the scalp surface. Notably, the simple contrasts between areas and montages showed deviations from planned electrode positions larger than 1 cm for the IFG, except for one spatial dimension. For M1, deviations were all smaller than 1 cm. Note that (Woods et al., 2015)) showed that a drift of 1-1.5 cm significantly altered the distribution of the electrical field.

### A.2. Electrical field simulation

*A.2.1. magnitude E*

*A.2.1.1. LMM for simulation differences of the magnitude E*

Detailed results of the linear mixed model (LMM) analysis for the dependent variable magnitude E are shown in **Table A.4**, post-hoc tests of main effects and simple contrasts are provided in **Table A.5** and **Figure 4**. Note that only simple effects and simple contrasts as interactions with the reference model are shown in the LMMs. The reference model of the LMM is the conventional IFG montage with the planned electrode position and a ROI radius of 1.25 cm that showed a mean magnitude of 0.14 V/m 95%-CI:[0.13, 0.15]. To simplify the interpretation, post-hoc analysis of main and simple effects as well as simple contrasts were calculated (**Figure 4** and **Table A.5**).

*A.2.2 Normal component of the electrical field (nE)*

*A.2.2.1. Simulation results of nE*

The normal component of the electrical field (nE) is visualized for both conventional and focal montages for planned (**Figures A.5A-D)** and actual (**Figure A.5E-H**) electrode positions. Positive and negative values are displayed for illustrative purposes in red (positive) and blue (negative). Note: Only positive values of nE were used for the ROI analyses (Antonenko et al., 2019).

As expected, higher field strengths across the entire brain were observed for conventional compared to focal montages for planned electrode positions ($95^{th}$ percentile mean±SD V/m conventional/focal: IFG 0.040±0.004/0.019±0.0004, t(58)=27.36, p<0.001; M1 0.043±0.005/ 0.016±0.004, t(58)=31.59, p<0.001). A similar pattern was confirmed for actual electrode positions (conventional/focal IFG: 0.040±0.001/ 0.016±0.003, t(58)=58.46, p<0.001, M1: 0.041±0.0001/ 0.014±0.003, t(58)=68.30, p<0.001).

*A.2.2.2. LMM for simulation differences of nE*

The LMM used for data analysis was identical to the one reported for magnitude E. Results of the LMM for different montages, areas, electrode positions and ROI radii are detailed in **Table A.6**. Estimated marginal means (EMMs) of fixed effects from the LMM model were calculated to compare main effects and simple contrasts, details are reported in **Table A.7**.

*A.2.2.3. EMMs of LMM for simulation differences of nE*

In line with the magnitude E analysis, post-hoc tests for main effects showed no differences between the two target areas for nE (IFG-M1: 0 V/m, 95%-CI: [0, 0.01]; t(115)=0.518; p=0.606; **Figure A.6A**). The most pronounced significant difference in simulated current flow was found when comparing ROI radii (r=1.25-r=2.5: 0.02 V/m 95%-CI: [0.01 0.02]; t(826)=15.76, p<0.001; **Figure A.6C)**, showing a decrease of nE for the larger ROI. Significant decreases of nE were also found for actual vs. planned electrode positions (0.01 V/m 95%-CI: [0.012, 0.014]; t(826) =24.84, p<0.001; **Figure A.6B)** and focal vs. conventional montages (0.01 V/m 95%-CI: [0, 0.02]; t(115)=3.802, p<0.001; **Figure A.6D)**. Therefore, these small changes in nE were more dependent on ROI radius, compared to the magnitude E analysis, where differences between montages were more pronounced (**Figure 4D)**. However, the direction of effects was identical for both parameters.

Electrode positioning errors (planned, actual) significantly decreased nE for the focal montages in both ROIs (**Figure A.6E**, lower panels: r=1.25, 0.03 V/m 95%-CI: [0.03, 0.03]; t(826)=22.84, p<0.001; r=2.5, 0.02 V/m, 95%-CI: [0.01, 0.02]; t(826)=11.72, p<0.001). This corresponds to a dose reduction of ~30% for both ROIs. In contrast, positioning errors did not affect nE for the conventional montages (**Figure A.6E**, upper panels: r=1.25: 0 V/m, 95%-CI: [0, 0.001]; t(826) =0.76, p=0.45; r=2.5: 0 V/m, 95%-CI: [-0.01, 0]; t(826)=-0.43, p=0.668). This is consistent with results from the magnitude E analysis.

Direct comparison of the two different ROI radii (1.25 cm, 2.5 cm) allowed further assessment of focality and precision of the stimulation. Increasing the ROI radius resulted only in a small reduction of nE for the conventional montages for both the planned and actual positions (**Figure A.6F,** upper panels; planned: 0.01 V/m, 95%-CI: [0.01, 0.01]; t(826)=6,32, p<0.001; actual: 0.01 V/m, 95%-CI: [0, 0.01]; t(826)=4.97, p<0.001). Significant decreases of nE in the larger compared to the smaller ROI were found for planned and actual positions, with larger effects for planned positions (**Figure A.6F,** lower panels; planned: 0.03 V/m, 95%-CI: [0.03, 0.03]; t(826)=21.14, p<0.001; actual: 0.01 V/m, 95%-CI: [0.01, 0.02]; t(826)=9.38, p<0.001). The latter was explained by overall lower nE values in both ROIs for actual electrode positions (see **Table A.7**).

Importantly, with *planned positioning* of the electrodes, focal montages resulted in significantly higher nE in the immediate target region (r=1.25 cm) compared to the conventional montages (**Figure A.6G**, top left panel; r=1.25: -0.01 V/m, 95%-CI: [-0.03, -0.01]; t(151)=-4.61, p<0.001), corresponding to a dose increase of 12.5%. This effect is mostly driven by the IFG (**Table A.7**). Note: The direction of results is identical compared to nE reported in the main part of the manuscript, even though there was only a trend for higher magnitude E (**Figure 4G**).

The higher precision of the focal compared to conventional set-ups is illustrated by a significant decrease of nE in the larger ROI (**Figure A.6G,** top right; 0.01 V/m, 95%-CI: [0.01, 0.03]; t(151)=2.76, p=0.01). Most importantly, deviations from the planned electrode positions (i.e., *actual positions*) resulted in a significant decrease of nE for focal compared to conventional montages for both ROI radii (**Figure A.6G,** bottom left; r=1.25: 0.02 V/m, 95%-CI: [0.01, 0.03]; t(151)=6.93, p<0.001; bottom right: r=2.5, 0.03 V/m, 95%-CI: [0.03, 0.02]; t(151)=9.1, p<0.001). This corresponds to a dose reduction of 25% and 42.86% respectively.

Details of the LLM analysis for the dependent variable nE are shown in **Table A.6**. The reference model was identical as for the magnitude E analysis (see **Table A.4**).

#### A.3 Focality of focal and conventional montages

In the main text, the precision of the stimulation was investigated using two overlapping ROIs (r=1.25; r=2.5 cm; see **Figure 4F** and **Figure A.6F**). Here, we present an additional analysis that directly addresses focality by comparing E-fields in the smaller ROI (r=1.25, inner sphere) with the larger ROI (r=2.5 cm), but the latter without the inner sphere (outer sphere). Details of the results are displayed in **Figure A.7**. Main effects for focal and conventional montages across all areas and electrode positions revealed an highly significant reduction of mean magnitude E in the outer sphere (0.099±0.03 V/m) compared to the inner sphere (0.130±0.04 V/m, t(239)=9.03, p<0.001) for focal montages. A significant, but much less pronounced reduction of magnitude E in the outer sphere was also found for the conventional montages (inner sphere: 0.142±0.02 V/m; outer sphere: 0.138±0.02, t(239)=2.43, p=0.02). Magnitude E reductions in the outer sphere were significantly more pronounced for focal compared to conventional montages (interaction sphere x montage -0.026±0.02 V/m, t(479)=-12.32, p<0.001). The normal component of the E-field showed a similar pattern. A more pronounced decrease was found for the focal (inner sphere: 0.072±0.03, outer sphere: 0.051±0.02, t(239)=9.43, p<0.001) compared to the conventional montages (inner sphere: 0.076±0.02, outer sphere: 0.068±0.01; t(239)=6.55, p<0.001; interaction sphere x montage: -0.01±0.01 V/m, t(479)=-7.348, p<0.001) montages. Therefore, we confirmed that the overall decrease of E-fields outside the immediate target region (r=1.25) was more pronounced for focal compared to conventional montages, indicating higher focality of current delivery for these set-ups.

#### A.4. References

Woods AJ, Bryant V, Sacchetti D, Gervits F, Hamilton R. Effects of Electrode Drift in Transcranial Direct Current Stimulation. Brain Stimul 2015;8:515–9. https://doi.org/10.1016/j.brs.2014.12.007.

Antonenko D, Thielscher A, Saturnino GB, Aydin S, Ittermann B, Grittner U, et al. Towards precise brain stimulation: Is electric field simulation related to neuromodulation? Brain Stimul. 2019b;12:1159–68. https://doi.org/10.1016/j.brs.2019.03.072
