## Appendix Tables for "Electrode positioning errors reduce current dose for focal tDCS set-ups: Evidence from individualized electric field mapping"

**Appendix Table A.1**: MNI coordinates for scalp positions and orientations of planned electrode configurations.

| Montage | | Anode | Cathode 1 | Cathode 2 | Cathode 3 |
|---|---|---|---|---|---|
| focal | IFG | -69/35/17 | -72/32 /-28 | -75/-1/41 | -40/66/33 |
| focal | M1 | -68/-13/62 | -33/-11/90 | -71/20/33 | -75/-54/46 |
| | | Anode | Cathode 1 | Orientation Anode | Orientation Cathode |
| conventional | IFG | -69/35/17 | 37/76/21 | -80/-6/21 | 31/54/56 |
| conventional | M1 | -68/-13/62 | 37/76/21 | -59/43/49 | 31/54/56 |

*MNI coordinates for scalp positions and orientations of planned electrode configurations. Conventional montages used the same positions for the cathode.IFG=inferior frontal gyrus; M1=primary motor cortex*

**Appendix Table A.2**: Results of linear mixed model analysis for Coordinate difference

| | Diff in \|mm\| | | | | |
|---|---|---|---|---|---|
| *Predictors*<br>*Factor [level]* × Interaction | *Estimates* | *95%-CI* | *p-value* | *df* | $R^2$ |
| (Intercept) | 14.92 | 13.09 – 15.50 | **<0.001** | 1423 | 0.289 |
| Sex [männlich] | -1.20 | -1.95 – -0.45 | **0.002** | 1423 | 0.013 |
| Treatment [Sham] | -0.02 | -0.77 – 0.73 | 0.968 | 1423 | 0 |
| Session [2] | -0.13 | -0.88 – 0.62 | 0.727 | 1423 | 0 |
| **ref: IFG conventional y dymension** | | | | | |
| Area [M1] | -4.64 | -6.06 – -3.21 | **<0.001** | 1423 | 0.033 |
| Montage [focal] | -2.63 | -4.06 – -1.21 | **<0.001** | 1423 | 0.011 |
| Dimension [z] | -2.29 | -3.46 – -1.12 | **<0.001** | 1423 | 0.008 |
| Dimension [x] | -3.43 | -4.60 – -2.26 | **<0.001** | 1423 | 0.018 |
| Area [M1] × Montage [focal] | 2.57 | 0.56 – 4.59 | **0.012** | 1423 | 0.005 |
| Area [M1] × Dimension [z] | -1.32 | -2.97 – 0.34 | 0.118 | 1423 | 0.001 |
| Area [M1] × Dimension [x] | -1.42 | -3.08 – 0.23 | 0.092 | 1423 | 0.002 |
| Montage [focal] × Dimension [z] | 5.24 | 3.59 – 6.90 | **<0.001** | 1423 | 0.021 |
| Montage [focal] × Dimension [x] | -1.29 | -2.94 – 0.37 | 0.127 | 1423 | 0.001 |
| (Area [M1] × Montage [focal]) × Dimension [z] | -2.41 | -4.75 – -0.07 | **0.044** | 1423 | 0.002 |
| (Area [M1] × Montage [focal]) × Dimension [x] | 1.19 | -1.15 – 3.53 | 0.319 | 1423 | 0.001 |
| Random Effects | | | | | |
| $\sigma^2$ | 21.36 | | | | |
| $\tau_{00}$ lmm_Subjects | 515 | | | | |
| ICC | 0.19 | | | | |
| N lmm_group | 240 | | | | |
| Observations | 1440 | | | | |

| | |
|---|---|
| Marginal $R^2$ / Conditional $R^2$ | 0.289 / 0.427 |

*Parameter of fixed and random effects of the linear mixed model for dependent variable coordinate difference and fixed effects Montage (conventional, focal), Area (IFG, M1), Electrode dimension x/y/z. Reference model (ref) conventional montage at area IFG for y dimension. Regression coefficients of linear mixed models (random intercept models) and two-sided p-values are reported. Semi-partial R² statistic as measure of effect size, approximated with Nakagawa and Schielzeth approach. CI=confidence interval. IFG=inferior frontal gyrus; M1=primary motor cortex, r= radius, df= degress of freedom,* $\sigma^2$ pooled residual variance, τ pooled variance explained by subjects, ICC=*Intraclass correlation coefficient* .

**Appendix Table A.3**: Posthoc estimated marginal means of linear mixed model analysis of Coordinates differences

| Posthoc EMMs of LMM analysis Coordinates differences | | | | | | | | | |
|---|---|---|---|---|---|---|---|---|---|
| Montage | Area | Electr. | contrast | EMMs | Lower. 95%-CI | Upper. 95%-CI | df | t-ratio | p-value |
| | | | | **Main Effects** | | | | | |
| conv | | | | 8.93 | 8.28 | 9.58 | 233 | | |
| focal | | | | 8.7 | 8.05 | 9.35 | 233 | | |
| | | | conv - focal | 0.23 | −0.69 | 1.15 | 233 | 0.60 | 0.55 |
| | IFG | | | 11.05 | 10.4 | 11.7 | 233 | | |
| | M1 | | | 6.59 | 5.93 | 7.24 | 233 | | |
| | | | IFG - M1 | 4.47 | 3.55 | 5.39 | 233 | 11.71 | <0.001 |
| | | y | | 10.63 | 9.95 | 11.31 | 681 | | |
| | | z | | 9.69 | 9.01 | 10.37 | 681 | | |
| | | x | | 6.14 | 5.46 | 6.82 | 681 | | |
| | | | y - z | 0.93 | 0.15 | 1.72 | 1192 | 3.13 | 0.01 |
| | | | y - x | 4.49 | 3.7 | 5.28 | 1192 | 15.05 | <0.001 |
| | | | z - x | 3.56 | 2.77 | 4.34 | 1192 | 11.92 | <0.001 |
| | | | | **Simple Contrasts** | | | | | |
| conv | IFG | y | | 13.62 | 12.03 | 15.21 | 681 | | |
| conv | IFG | z | | 11.32 | 9.74 | 12.91 | 681 | | |
| conv | IFG | x | | 10.19 | 8.6 | 11.77 | 681 | | |
| conv | M1 | y | | 8.98 | 7.39 | 10.57 | 680 | | |
| conv | M1 | z | | 5.37 | 3.78 | 6.96 | 680 | | |
| conv | M1 | x | | 4.13 | 2.54 | 5.71 | 680 | | |
| focal | IFG | y | | 10.99 | 9.4 | 12.57 | 680 | | |
| focal | IFG | z | | 13.93 | 12.35 | 15.52 | 680 | | |
| focal | IFG | x | | 6.27 | 4.68 | 7.85 | 680 | | |
| focal | M1 | y | | 8.92 | 7.33 | 10.51 | 680 | | |
| focal | M1 | z | | 8.14 | 6.56 | 9.73 | 680 | | |
| focal | M1 | x | | 3.97 | 2.38 | 5.56 | 680 | | |
| conv | IFG | | y - z | 2.29 | 0.45 | 4.13 | 1192 | 3.84 | <0.001 |
| conv | IFG | | y - x | 3.43 | 1.59 | 5.27 | 1192 | 5.75 | <0.001 |
| conv | IFG | | z - x | 1.14 | −0.7 | 2.98 | 1192 | 1.91 | 0.14 |
| conv | M1 | | y - z | 3.61 | 1.77 | 5.45 | 1192 | 6.05 | <0.001 |

| | | | | | | | | | |
|---|---|---|---|---|---|---|---|---|---|
| conv | M1 | | y - x | 4.86 | 3.01 | 6.7 | 1192 | 8.14 | <0.001 |
| conv | M1 | | z - x | 1.24 | −0.6 | 3.08 | 1192 | 2.08 | 0.09 |
| focal | IFG | | y - z | −2.95 | −4.79 | −1.11 | 1192 | −4.94 | <0.001 |
| focal | IFG | | y - x | 4.72 | 2.88 | 6.56 | 1192 | 7.91 | <0.001 |
| focal | IFG | | z - x | 7.67 | 5.83 | 9.51 | 1192 | 12.85 | <0.001 |
| focal | M1 | | y - z | 0.78 | −1.06 | 2.62 | 1192 | 1.30 | 0.39 |
| focal | M1 | | y - x | 4.95 | 3.11 | 6.79 | 1192 | 8.30 | <0.001 |
| focal | M1 | | z - x | 4.18 | 2.33 | 6.02 | 1192 | 7.00 | <0.001 |
| conv | IFG | y | | 13.62 | 12.08 | 15.16 | 681 | | |
| conv | M1 | y | | 8.98 | 7.44 | 10.52 | 680 | | |
| conv | IFG | z | | 11.32 | 9.78 | 12.87 | 681 | | |
| conv | M1 | z | | 5.37 | 3.83 | 6.91 | 680 | | |
| conv | IFG | x | | 10.19 | 8.64 | 11.73 | 681 | | |
| conv | M1 | x | | 4.13 | 2.58 | 5.67 | 680 | | |
| focal | IFG | y | | 10.99 | 9.44 | 12.53 | 680 | | |
| focal | M1 | y | | 8.92 | 7.38 | 10.46 | 680 | | |
| focal | IFG | z | | 13.93 | 12.39 | 15.48 | 680 | | |
| focal | M1 | z | | 8.14 | 6.6 | 9.69 | 680 | | |
| focal | IFG | x | | 6.27 | 4.72 | 7.81 | 680 | | |
| focal | M1 | x | | 3.97 | 2.43 | 5.51 | 680 | | |
| conv | | y | IFG - M1 | 4.64 | 2.46 | 6.82 | 681 | 6.38 | <0.001 |
| conv | | z | IFG - M1 | 5.96 | 3.77 | 8.14 | 681 | 8.20 | <0.001 |
| conv | | x | IFG - M1 | 6.06 | 3.88 | 8.24 | 681 | 8.34 | <0.001 |
| focal | | y | IFG - M1 | 2.06 | −0.12 | 4.25 | 680 | 2.84 | 0.01 |
| focal | | z | IFG - M1 | 5.79 | 3.61 | 7.97 | 680 | 7.96 | <0.001 |
| focal | | x | IFG - M1 | 2.30 | 0.12 | 4.48 | 680 | 3.16 | <0.001 |
| y | IFG | conv | | 13.62 | 12.08 | 15.16 | 681 | | |
| y | IFG | focal | | 10.99 | 9.44 | 12.53 | 680 | | |
| z | IFG | conv | | 11.32 | 9.78 | 12.87 | 681 | | |
| z | IFG | focal | | 13.93 | 12.39 | 15.48 | 680 | | |
| x | IFG | conv | | 10.19 | 8.65 | 11.73 | 681 | | |
| x | IFG | focal | | 6.27 | 4.72 | 7.81 | 680 | | |
| y | M1 | conv | | 8.98 | 7.44 | 10.52 | 680 | | |
| y | M1 | focal | | 8.92 | 7.38 | 10.47 | 680 | | |
| z | M1 | conv | | 5.37 | 3.83 | 6.91 | 680 | | |
| z | M1 | focal | | 8.15 | 6.6 | 9.69 | 680 | | |
| x | M1 | conv | | 4.13 | 2.58 | 5.67 | 680 | | |
| x | M1 | focal | | 3.97 | 2.43 | 5.51 | 680 | | |
| y | IFG | | conv - focal | 2.63 | 0.45 | 4.81 | 681 | 3.62 | <0.001 |
| z | IFG | | conv - focal | −2.61 | −4.791 | −0.429 | 681 | −3.59 | <0.001 |
| x | IFG | | conv - focal | 3.92 | 1.74 | 6.1 | 681 | 5.40 | <0.001 |
| y | M1 | | conv - focal | 0.06 | −2.122 | 2.24 | 681 | 0..81 | 0.94 |
| z | M1 | | conv - focal | −2.777 | −4.958 | −0.596 | 681 | −3.82 | <0.001 |
| x | M1 | | conv - focal | 0.16 | −2.025 | 2.34 | 681 | 0.22 | 0.83 |

*Posthoc test of the estimated marginal means (EMMs) of main effects and simple contrasts for the linear mixed model (LMM) of coordinate difference. Level of main factors are given in the first 3 rcolumns. Direct comparison between two level is given in the contrast column. Level in the same row as contrasts are the within level of the second and third factor for the contrast. Posthoc test statistic values of contrast between the EMMs and associates lower and upper part of the 95%*

*Confidence intervals (95%-CI) are given in the last 3 rows with degrees of freedom (df), t-test statistic (t-statistic) and p-values (p). All tests are corrected for multiple comparisons with the Tukey method.*

**Appendix Table A.4**: Results of linear mixed model analysis for magnitude of electrical field strength E (magnitude E)

| | meanE | | | | |
|---|---|---|---|---|---|
| Predictors | Estimates | 95%-CI | p-value | df | $R^2$ |
| (Intercept) | 0.14 | 0.13 – 0.15 | **<0.001** | 130 | |
| Sex [male] | -0.01 | -0.01 – 0.00 | 0.08 | 115 | 0.01 |
| Session [2] | -0.00 | -0.00 – 0.00 | 0.39 | 594 | |
| **ref: conventional ROI radius 1.25 cm planned IFG** | | | | | |
| Treatment [Sham] | 0.00 | -0.00 – 0.00 | 0.71 | 594 | |
| Montage [focal] | 0.02 | 0.00 – 0.04 | **0.01** | 117 | 0.02 |
| ROI radius [r=2.5] | -0.00 | -0.01 – 0.00 | 0.27 | 175 | 0 |
| Electrode position[actual] | 0.01 | 0.01 – 0.02 | **<0.001** | 154 | 0.01 |
| Area [M1] | 0.01 | -0.01 – 0.03 | 0.29 | 117 | 0 |
| Montage [focal] × ROI radius [r=2.5] | -0.04 | -0.04 – -0.03 | **<0.001** | 175 | 0.03 |
| Montage [focal] × Electrode position [actual] | -0.06 | -0.07 – -0.05 | **<0.001** | 154 | 0.07 |
| ROI radius [r=2.5] × Electrode position [actual] | 0.00 | -0.00 – 0.01 | 0.55 | 594 | 0 |
| Montage [focal] × Area [M1] | -0.02 | -0.05 – 0.00 | 0.06 | 117 | 0.01 |
| ROI radius [r=2.5] × Area [M1] | -0.00 | -0.01 – 0.00 | 0.14 | 175 | 0 |
| Electrode position [actual] × Area [M1] | -0.02 | -0.03 – -0.01 | **<0.001** | 154 | 0.01 |
| (Montage [focal] × ROI radius [r=2.5]) × Electrode position [actual] | 0.01 | 0.01 – 0.02 | **<0.001** | 594 | 0 |
| (Montage [focal] × ROI radius [r=2.5]) × Area [M1] | 0.01 | 0.00 – 0.02 | **0.04** | 175 | 0 |
| (Montage [focal] × Electrode position [actual]) × Area [M1] | 0.02 | 0.01 – 0.04 | **<0.001** | 154 | 0.01 |
| (ROI radius [r=2.5] × | -0.00 | -0.01 – 0.00 | 0.75 | 594 | 0 |

| | | | | | |
|---|---|---|---|---|---|
| Electrode position [actual]) × Area [M1] | | | | | |
| (Montage [focal] × ROI radius [r=2.5] × Electrode position [actual]) × Area [M1] | 0.00 | -0.00 – 0.01 | 0.44 | 594 | 0 |
| Random Effects | | | | | |
| $\sigma^2$ | 0.00 | | | | |
| $\tau_{00}$ lmm_Subjects | 0.00 | | | | |
| $\tau_{11}$ lmm_Subjects.ROI radius [r=2.5] | 0.00 | | | | |
| $\tau_{11}$ lmm_Subjects.Electrode position [actual] | 0.00 | | | | |
| $\rho_{01}$ | -0.83 | | | | |
| | -0.62 | | | | |
| ICC | 0.91 | | | | |
| N lmm_Subjects | 120 | | | | |
| Observations | 960 | | | | |
| Marginal $R^2$ / Conditional $R^2$ | 0.400 / 0.949 | | | | |

*Parameter of fixed and random effects of the linear mixed model for dependent variable magnitude E and fixed effects Montage (conventional, focal), Area (IFG, M1), Electrode position (planned, actual) and ROI radius (r=1.25, r=2.5). Reference model (ref) conventional montage at area IFG for planned electrode position with ROI radius of 1.25 cm. Regression coefficients of linear mixed models (random intercept models) and two-sided p-values are reported. Semi-partial R² statistic as measure of effect size, approximated with Nakagawa and Schielzeth approach. CI=confidence interval. IFG=inferior frontal gyrus; M1=primary motor cortex, r= radius, df= degress of freedom,* $\sigma^2$ pooled residual variance, $\tau_{00}$ pooled variance explained by subjects, $\tau_{11}$ pooled variance explained by subjects and by factor ROI radius or eelctrode position, ρ = correlation coefficient, ICC=*Intraclass correlation coefficient, N= total number of subjects.*

**Appendix Table A.5**: Posthoc estimated marginal means of linear mixed model analysis of magnitude E

| Posthoc EMMs of LMM magnitude E | | | | | | | | | | |
|---|---|---|---|---|---|---|---|---|---|---|
| Montage | Area | Position | ROI radius | Contrast | EMMs | Lower. 95%-CI | Upper. 95%-CI | df | t-statistic | p-value |
| | | | | | **Main Effects** | | | | | |
| conv | | | | | 0.14 | 0.13 | 0.15 | 115 | | |
| focal | | | | | 0.11 | 0.11 | 0.12 | 115 | | |
| | | | | conv - focal | 0.03 | 0.02 | 0.04 | 115 | 5.68 | <0.001 |

| | | | | | | | | | | |
|---|---|---|---|---|---|---|---|---|---|---|
| | IFG | | | | 0.13 | 0.12 | 0.14 | 115 | | |
| | M1 | | | | 0.12 | 0.12 | 0.13 | 115 | | |
| | | | | IFG - M1 | 0.01 | −0.006 | 0.02 | 115 | 1.12 | 0.263 |
| | | planned | | | 0.14 | 0.13 | 0.14 | 122 | | |
| | | actual | | | 0.12 | 0.11 | 0.12 | 122 | | |
| | | | | plan - actual | 0.02 | 0.01 | 0.02 | 826 | 12.56 | <0.001 |
| | | | 1.25 | | 0.14 | 0.13 | 0.14 | 122 | | |
| | | | 2.5 | | 0.12 | 0.11 | 0.12 | 122 | | |
| | | | | 1.25 – 2.5 | 0.02 | 0.02 | 0.02 | 826 | 16.96 | <0.001 |
| | | | | **Simple Contrasts** | | | | | | |
| conv | | planned | 1.25 | | 0.14 | 0.13 | 0.15 | 136 | | |
| conv | | actual | 1.25 | | 0.14 | 0.13 | 0.15 | 136 | | |
| focal | | planned | 1.25 | | 0.15 | 0.14 | 0.16 | 136 | | |
| focal | | actual | 1.25 | | 0.11 | 0.1 | 0.12 | 136 | | |
| conv | | planned | 2.5 | | 0.14 | 0.13 | 0.15 | 136 | | |
| conv | | actual | 2.5 | | 0.14 | 0.13 | 0.15 | 136 | | |
| focal | | planned | 2.5 | | 0.11 | 0.1 | 0.12 | 136 | | |
| focal | | actual | 2.5 | | 0.08 | 0.08 | 0.09 | 136 | | |
| conv | | | 1.25 | plan - actual | −0.003 | -0.01 | 0 | 826 | −1.99 | 0.127 |
| focal | | | 1.25 | plan - actual | 0.04 | 0.04 | 0.05 | 826 | 27.99 | <0.001 |
| conv | | | 2.5 | plan - actual | −0.004 | -0.01 | 0 | 826 | −2.50 | 0.056 |
| focal | | | 2.5 | plan - actual | 0.03 | 0.02 | 0.04 | 826 | 19.4 | <0.001 |
| conv | | planned | 1.25 | | 0.14 | 0.13 | 0.15 | 136 | | |
| conv | | planned | 2.5 | | 0.14 | 0.13 | 0.15 | 136 | | |
| focal | | planned | 1.25 | | 0.15 | 0.14 | 0.16 | 136 | | |
| focal | | planned | 2.5 | | 0.11 | 0.1 | 0.12 | 136 | | |
| conv | | actual | 1.25 | | 0.14 | 0.13 | 0.15 | 136 | | |
| conv | | actual | 2.5 | | 0.14 | 0.13 | 0.15 | 136 | | |
| focal | | actual | 1.25 | | 0.11 | 0.1 | 0.12 | 136 | | |
| focal | | actual | 2.5 | | 0.08 | 0.08 | 0.09 | 136 | | |
| conv | | planned | | 1.25 - 2.5 | 0.01 | 0 | 0.01 | 826 | 3.32 | 0.003 |
| focal | | planned | | 1.25 - 2.5 | 0.04 | 0.03 | 0.04 | 826 | 24.62 | <0.001 |
| conv | | actual | | 1.25 - 2.5 | 0.004 | −0.001 | 0.01 | 826 | 2.81 | 0.010 |
| focal | | actual | | 1.25 - 2.5 | 0.02 | 0.02 | 0.03 | 826 | 16.02 | <0.001 |
| conv | | planned | 1.25 | | 0.14 | 0.13 | 0.15 | 136 | | |
| focal | | planned | 1.25 | | 0.15 | 0.14 | 0.16 | 136 | | |
| conv | | actual | 1.25 | | 0.14 | 0.13 | 0.15 | 136 | | |
| focal | | actual | 1.25 | | 0.11 | 0.1 | 0.12 | 136 | | |
| conv | | planned | 2.5 | | 0.14 | 0.13 | 0.15 | 136 | | |
| focal | | planned | 2.5 | | 0.11 | 0.1 | 0.12 | 136 | | |
| conv | | actual | 2.5 | | 0.14 | 0.13 | 0.15 | 136 | | |
| focal | | actual | 2.5 | | 0.08 | 0.08 | 0.09 | 136 | | |
| | | planned | 1.25 | conv - focal | −0.010 | −0.028 | 0.01 | 136 | −1.73 | 0.09 |
| | | actual | 1.25 | conv - focal | 0.04 | 0.02 | 0.05 | 136 | 7.22 | <0.001 |
| | | planned | 2.5 | conv - focal | 0.02 | 0.01 | 0.04 | 136 | 4.81 | <0.001 |
| | | actual | 2.5 | conv - focal | 0.06 | 0.05 | 0.07 | 136 | 15.43 | <0.001 |
| conv | IFG | planned | 1.25 | | 0.14 | 0.12 | 0.16 | 136 | | |

| | | | | | | | | | | |
|---|---|---|---|---|---|---|---|---|---|---|
| focal | IFG | planned | 1.25 | | 0.16 | 0.14 | 0.18 | 136 | | |
| conv | IFG | actual | 1.25 | | 0.15 | 0.13 | 0.16 | 136 | | |
| focal | IFG | actual | 1.25 | | 0.11 | 0.1 | 0.13 | 136 | | |
| conv | IFG | planned | 2.5 | | 0.13 | 0.12 | 0.15 | 136 | | |
| focal | IFG | planned | 2.5 | | 0.12 | 0.1 | 0.13 | 136 | | |
| conv | IFG | actual | 2.5 | | 0.15 | 0.13 | 0.16 | 136 | | |
| focal | IFG | actual | 2.5 | | 0.08 | 0.07 | 0.1 | 136 | | |
| conv | M1 | planned | 1.25 | | 0.15 | 0.13 | 0.16 | 136 | | |
| focal | M1 | planned | 1.25 | | 0.14 | 0.13 | 0.16 | 136 | | |
| conv | M1 | actual | 1.25 | | 0.14 | 0.12 | 0.16 | 136 | | |
| focal | M1 | actual | 1.25 | | 0.11 | 0.09 | 0.12 | 136 | | |
| conv | M1 | planned | 2.5 | | 0.14 | 0.12 | 0.15 | 136 | | |
| focal | M1 | planned | 2.5 | | 0.11 | 0.09 | 0.12 | 136 | | |
| conv | M1 | actual | 2.5 | | 0.13 | 0.12 | 0.14 | 136 | | |
| focal | M1 | actual | 2.5 | | 0.08 | 0.07 | 0.1 | 136 | | |
| | IFG | planned | 1.25 | conv - focal | −0.022 | −0.048 | 0.01 | 136 | −2.57 | 0.01 |
| | IFG | actual | 1.25 | conv - focal | 0.04 | 0.02 | 0.06 | 136 | 5.26 | <0.001 |
| | IFG | planned | 2.5 | conv - focal | 0.02 | −0.0054 | 0.04 | 136 | 2.39 | 0.02 |
| | IFG | actual | 2.5 | conv - focal | 0.06 | 0.05 | 0.08 | 136 | 12.37 | <0.001 |
| | M1 | planned | 1.25 | conv - focal | 0 | −0.026 | 0.03 | 136 | 0.12 | 0.9 |
| | M1 | actual | 1.25 | conv - focal | 0.03 | 0.01 | 0.06 | 136 | 4.96 | <0.001 |
| | M1 | planned | 2.5 | conv - focal | 0.03 | 0.01 | 0.05 | 136 | 4.42 | <0.001 |
| | M1 | actual | 2.5 | conv - focal | 0.05 | 0.03 | 0.06 | 136 | 9.45 | <0.001 |

*Table for the posthoc test of the estimated marginal means (EMMs) of main effects and simple contrasts for the linear mixed model (LMM) of  electric field magnitude (magnitude E). Level of main factors are given in the first 4 columns. Direct comparison between two level is given in the contrast column. Level in the same row as contrasts are the within level of the second and third factor for the contrast. Posthoc test statistic values of contrast between the EMMs and associates lower and upper part of the 95% Confidence intervals (95%-CI) are given in the last 3 rows with degrees of freedom (df), t-test statistic (t-statistic) and p-values (p). All tests are corrected for multiple comparisons with the Tukey method.*

**Appendix Table A.6**: Results of linear mixed model analysis for normal component of the electrical field strength E

| **Mean E normal component** | | | | | |
|---|---|---|---|---|---|
| *Predictors* | *Estimates* | *95%-CI* | *p-value* | *df* | *$R^2$* |
| (Intercept) | 0.07 | 0.06 – 0.08 | **<0.001** | 127.06 | |
| Sex [male] | -0.00 | -0.00 – 0.00 | 0.54 | 115.00 | 0.001 |
| Session [2] | -0.00 | -0.00 – 0.00 | 0.12 | 594.00 | 0.000 |
| Treatment [Sham] | 0.00 | -0.00 – 0.00 | 0.73 | 594.00 | 0.000 |
| **ref: conventional ROI radius 1.25 cm planned IFG** |  |  |  |  |  |
| Montage [focal] | 0.03 | 0.02 – 0.04 | **<0.001** | 117.51 | 0.076 |
| ROI radius [r=2.5] | -0.00 | -0.01 – 0.00 | 0.11 | 151.24 | 0.001 |
| Electrode position [actual] | 0.01 | 0.00 – 0.01 | **0.02** | 146.91 | 0.003 |
| Area [M1] | 0.02 | 0.01 – 0.03 | **0.04** | 117.51 | 0.029 |
| Montage [focal] × ROI radius [r=2.5] | -0.03 | -0.03 – -0.02 | **<0.001** | 151.24 | 0.038 |
| Montage [focal] × Electrode position [actual] | -0.04 | -0.05 – -0.04 | **<0.001** | 146.91 | 0.088 |

| | | | | | |
|---|---|---|---|---|---|
| ROI radius [r=2.5] × Electrode position [actual] | 0.00 | -0.00 – 0.00 | 0.46 | 594.00 | 0.000 |
| Montage [focal] × Area [M1] | -0.03 | -0.04 – -0.01 | **<0.001** | 117.50 | 0.044 |
| ROI radius [r=2.5] × Area [M1] | -0.01 | -0.02 – -0.01 | **<0.001** | 151.24 | 0.007 |
| Electrode position [actual] × Area [M1] | -0.01 | -0.02 – -0.01 | **<0.001** | 146.91 | 0.009 |
| (Montage [focal] × ROI radius [r=2.5]) × Electrode position [actual] | 0.01 | 0.01 – 0.02 | **<0.001** | 594.00 | 0.005 |
| (Montage [focal] × ROI radius [r=2.5]) × Area [M1] | 0.01 | 0.00 – 0.02 | **0.03** | 151.24 | 0.004 |
| (Montage [focal] × Electrode position [actual] × Area [M1] | 0.02 | 0.01 – 0.03 | **<0.001** | 146.91 | 0.001 |
| (ROI radius [r=2.5] × Electrode position [actual] × Area [M1] | 0.00 | -0.00 – 0.01 | 0.47 | 594.00 | 0.000 |
| (Montage [focal] × ROI radius [r=2.5] × Electrode position [actual]) × Area [M1] | 0.00 | -0.00 – 0.01 | 0.31 | 594.00 | 0.000 |
| **Random Effects** | | | | | |
| $\sigma^2$ | 0.00 | | | | |
| $\tau_{00}$ lmm_group | 0.00 | | | | |
| $\tau_{11}$ lmm_group.ROI radius r=2.5 | 0.00 | | | | |
| $\tau_{11}$ lmm_group.electrodepositionactual | 0.00 | | | | |
| $\rho_{01}$ | -0.86 | | | | |
| | -0.66 | | | | |
| ICC | 0.90 | | | | |
| N lmm_group | 120 | | | | |
| Observations | 960 | | | | |
| Marginal $R^2$ / Conditional $R^2$ | 0.413 / 0.940 | | | | |

*Parameter of fixed and random effects of the linear mixed model for dependent variable normal component of the electrical field E (nE) and fixed effects Montage (conventional, focal), Area (IFG, M1), Electrode position (planned, actual) and ROI radius (r=1.25, r=2.5). Reference model (ref) conventional montage at area IFG for planned electrode position with ROI radius of 1.25 cm. Regression coefficients of linear mixed models (random intercept models) and two-sided p-values are reported. Semi-partial $R^2$ statistic as measure of effect size, approximated with Nakagawa and Schielzeth approach. CI=confidence interval. IFG=inferior frontal gyrus; M1=primary motor cortex, r= radius, df= degress of freedom,* $\sigma^2$ pooled residual variance, $\tau_{00}$ pooled variance explained by subjects, $\tau_{11}$ pooled variance explained by subjects and by factor ROI radius or eelctrode position, ρ = correlation coefficient, ICC=*Intraclass correlation coefficient, N= total number of subjects.*

**Appendix Table A.7**: Posthoc estimated marginal means of linear mixed model analysis of nE

| Posthoc EMMs nE | | | | | | | | | | |
|---|---|---|---|---|---|---|---|---|---|---|
| Montage | Area | Position | ROI radius | Contrast | EMMs | Lower. 95%-CI | Upper. 95%-CI | df | t-statistic | p-value |
| | | | | **Main Effects** | | | | | | |
| conv | | | | | 0.07 | 0.07 | 0.08 | 115 | | |
| focal | | | | | 0.06 | 0.06 | 0.07 | 115 | | |
| | | | | conv - focal | 0.01 | 0.00 | 0.02 | 115 | 3.8 | <0.001 |
| | IFG | | | | 0.07 | 0.06 | 0.07 | 115 | | |
| | M1 | | | | 0.07 | 0.06 | 0.07 | 115 | | |
| | | | | IFG - M1 | 0.00 | 0.00 | 0.01 | 115 | 0.52 | 0.606 |
| | | planned | | | 0.07 | 0.07 | 0.08 | 127 | | |
| | | actual | | | 0.06 | 0.06 | 0.06 | 127 | | |
| | | . | | plan - actual | 0.01 | 0.01 | 0.02 | 826 | 13.09 | <0.001 |
| | | | 1.25 | | 0.07 | 0.07 | 0.08 | 127 | | |
| | | | 2.5 | | 0.06 | 0.06 | 0.06 | 127 | | |
| | | | | 1.25 - 2.5 | 0.02 | 0.01 | 0.02 | 826 | 15.76 | <0.001 |
| | | | | **Simple Contrasts** | | | | | | |
| conv | | planned | 1.25 | | 0.08 | 0.07 | 0.09 | 151 | | |
| focal | | planned | 1.25 | | 0.09 | 0.08 | 0.10 | 151 | | |
| conv | | planned | 2.5 | | 0.07 | 0.06 | 0.07 | 151 | | |
| focal | | planned | 2.5 | | 0.06 | 0.05 | 0.07 | 151 | | |
| conv | | actual | 1.25 | | 0.08 | 0.07 | 0.08 | 151 | | |
| focal | | actual | 1.25 | | 0.06 | 0.05 | 0.06 | 151 | | |
| conv | | actual | 2.5 | | 0.07 | 0.06 | 0.07 | 151 | | |
| focal | | actual | 2.5 | | 0.04 | 0.04 | 0.05 | 151 | | |
| | | planned | 1.25 | conv - focal | -0.01 | -0.03 | -0.01 | 151 | −4.61 | <0.001 |
| | | planned | 2.5 | conv - focal | 0.01 | 0.00 | 0.02 | 151 | 2.76 | 0.01 |
| | | actual | 1.25 | conv - focal | 0.02 | 0.01 | 0.03 | 151 | 6.93 | <0.001 |
| | | actual | 2.5 | conv - focal | 0.03 | 0.02 | 0.03 | 151 | 9.11 | <0.001 |
| conv | | planned | 1.25 | | 0.08 | 0.07 | 0.09 | 151 | | |
| conv | | actual | 1.25 | | 0.08 | 0.07 | 0.08 | 151 | | |
| focal | | planned | 1.25 | | 0.09 | 0.08 | 0.10 | 151 | | |
| focal | | actual | 1.25 | | 0.06 | 0.05 | 0.06 | 151 | | |
| conv | | planned | 2.5 | | 0.07 | 0.06 | 0.07 | 151 | | |
| conv | | actual | 2.5 | | 0.07 | 0.07 | 0.07 | 151 | | |
| focal | | planned | 2.5 | | 0.06 | 0.05 | 0.07 | 151 | | |
| focal | | actual | 2.5 | | 0.04 | 0.04 | 0.05 | 151 | | |
| conv | | | 1.25 | plan - actual | 0.00 | 0.00 | 0.01 | 826 | 0.76 | 0.45 |
| focal | | | 1.25 | plan - actual | 0.03 | 0.03 | 0.04 | 826 | 22.84 | <0.001 |
| conv | | | 2.5 | plan - actual | 0.00 | -0.01 | 0.00 | 826 | −0.43 | 0.668 |
| focal | | | 2.5 | plan - actual | 0.02 | 0.01 | 0.02 | 826 | 11.72 | <0.001 |

| | | | | | | | | | | |
|---|---|---|---|---|---|---|---|---|---|---|
| conv | | planned | 1.25 | | 0.08 | 0.07 | 0.09 | 151 | | |
| conv | | planned | 2.5 | | 0.07 | 0.06 | 0.07 | 151 | | |
| focal | | planned | 1.25 | | 0.09 | 0.08 | 0.10 | 151 | | |
| focal | | planned | 2.5 | | 0.06 | 0.05 | 0.07 | 151 | | |
| conv | | actual | 1.25 | | 0.08 | 0.07 | 0.08 | 151 | | |
| conv | | actual | 2.5 | | 0.07 | 0.07 | 0.07 | 151 | | |
| focal | | actual | 1.25 | | 0.06 | 0.05 | 0.06 | 151 | | |
| focal | | actual | 2.5 | | 0.04 | 0.04 | 0.05 | 151 | | |
| conv | | planned | | 1.25 - 2.5 | 0.01 | 0.01 | 0.01 | 826 | 6.23 | <0.001 |
| focal | | planned | | 1.25 - 2.5 | 0.03 | 0.03 | 0.03 | 826 | 21.14 | <0.001 |
| conv | | actual | | 1.25 - 2.5 | 0.01 | 0.00 | 0.01 | 826 | 4.97 | <0.001 |
| focal | | actual | | 1.25 - 2.5 | 0.01 | 0.01 | 0.02 | 826 | 9.38 | <0.001 |
| conv | IFG | planned | 1.25 | | 0.07 | 0.06 | 0.77 | 117 | | |
| focal | IFG | planned | 1.25 | | 0.10 | 0.09 | 0.11 | 118 | | |
| conv | IFG | actual | 1.25 | | 0.07 | 0.06 | 0.82 | 118 | | |
| focal | IFG | actual | 1.25 | | 0.06 | 0.05 | 0.68 | 119 | | |
| conv | IFG | planned | 2.5 | | 0.07 | 0.06 | 0.74 | 119 | | |
| focal | IFG | planned | 2.5 | | 0.07 | 0.06 | 0.75 | 119 | | |
| conv | IFG | actual | 2.5 | | 0.07 | 0.06 | 0.80 | 123 | | |
| focal | IFG | actual | 2.5 | | 0.04 | 0.03 | 0.52 | 123 | | |
| conv | M1 | planned | 1.25 | | 0.08 | 0.08 | 0.93 | 118 | | |
| focal | M1 | planned | 1.25 | | 0.08 | 0.07 | 0.92 | 118 | | |
| conv | M1 | actual | 1.25 | | 0.08 | 0.07 | 0.86 | 119 | | |
| focal | M1 | actual | 1.25 | | 0.05 | 0.04 | 0.61 | 119 | | |
| conv | M1 | planned | 2.5 | | 0.07 | 0.06 | 0.79 | 119 | | |
| focal | M1 | planned | 2.5 | | 0.05 | 0.05 | 0.63 | 119 | | |
| conv | M1 | actual | 2.5 | | 0.07 | 0.06 | 0.74 | 123 | | |
| focal | M1 | actual | 2.5 | | 0.04 | 0.03 | 0.51 | 123 | | |
| | IFG | planned | 1.25 | conv - focal | -0.03 | -0.05 | -0.01 | 118 | −4.87 | <0.001 |
| | IFG | actual | 1.25 | conv - focal | 0.02 | 0.00 | 0.29 | 119 | 3.36 | 0.001 |
| | IFG | planned | 2.5 | conv - focal | 0.00 | -0.01 | 0.011 | 119 | −0.15 | 0.878 |
| | IFG | actual | 2.5 | conv - focal | 0.03 | 0.02 | 0.37 | 123 | 10.53 | <0.001 |
| | M1 | planned | 1.25 | conv - focal | 0.00 | -0.02 | 0.19 | 117 | 0.27 | 0.79 |
| | M1 | actual | 1.25 | conv - focal | 0.02 | 0.01 | 0.38 | 118 | 5.6 | <0.001 |
| | M1 | planned | 2.5 | conv - focal | 0.02 | 0.00 | 0.28 | 119 | 4.31 | <0.001 |
| | M1 | actual | 2.5 | conv - focal | 0.02 | 0.02 | 0.32 | 123 | 8.69 | <0.001 |

*Posthoc test of the estimated marginal means (EMMs) of main effects and simple contrasts for the linear mixed model (LMM) of  the electrical field normal component (nE). Level of Main factors are given in the first 4 columns. Direct comparison between two level is given in the contrast column. Level in the same row as contrasts are the within level of the second and third factor for the contrast. Posthoc test statistic values of contrast between the EMMs and associates lower and upper part of the 95% Confidence intervals (95%-CI) are given in the last 3 rows with degrees of freedom (df), t-test statistic (t-statistic) and p-values (p). All tests are corrected for multiple comparisons with the Tukey method.*
