## Supplementary figures and images for "Electrode positioning errors reduce current dose for focal tDCS set-ups: Evidence from individualized electric field mapping"

### Appendix Figure 1

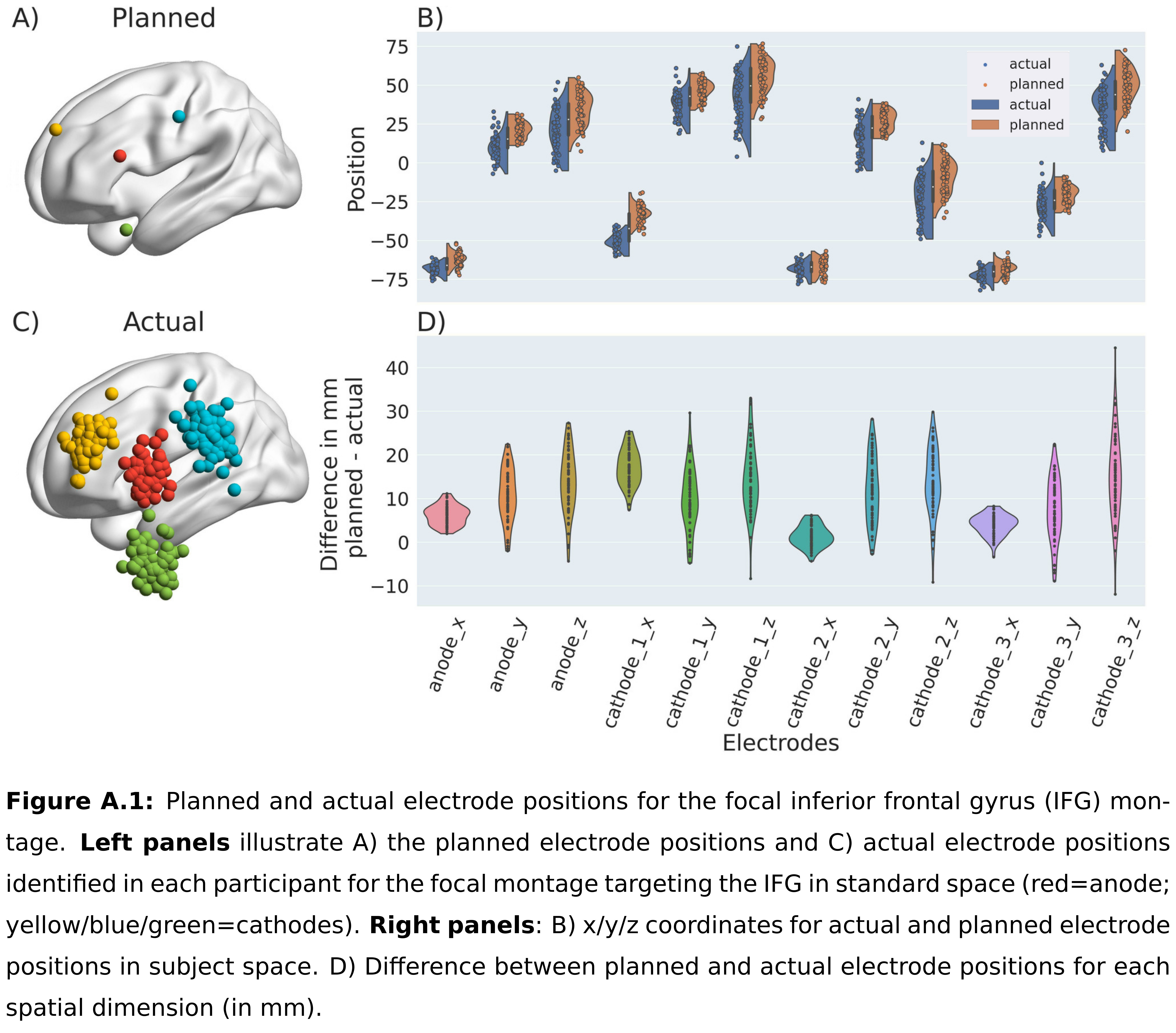

### Appendix Figure 2

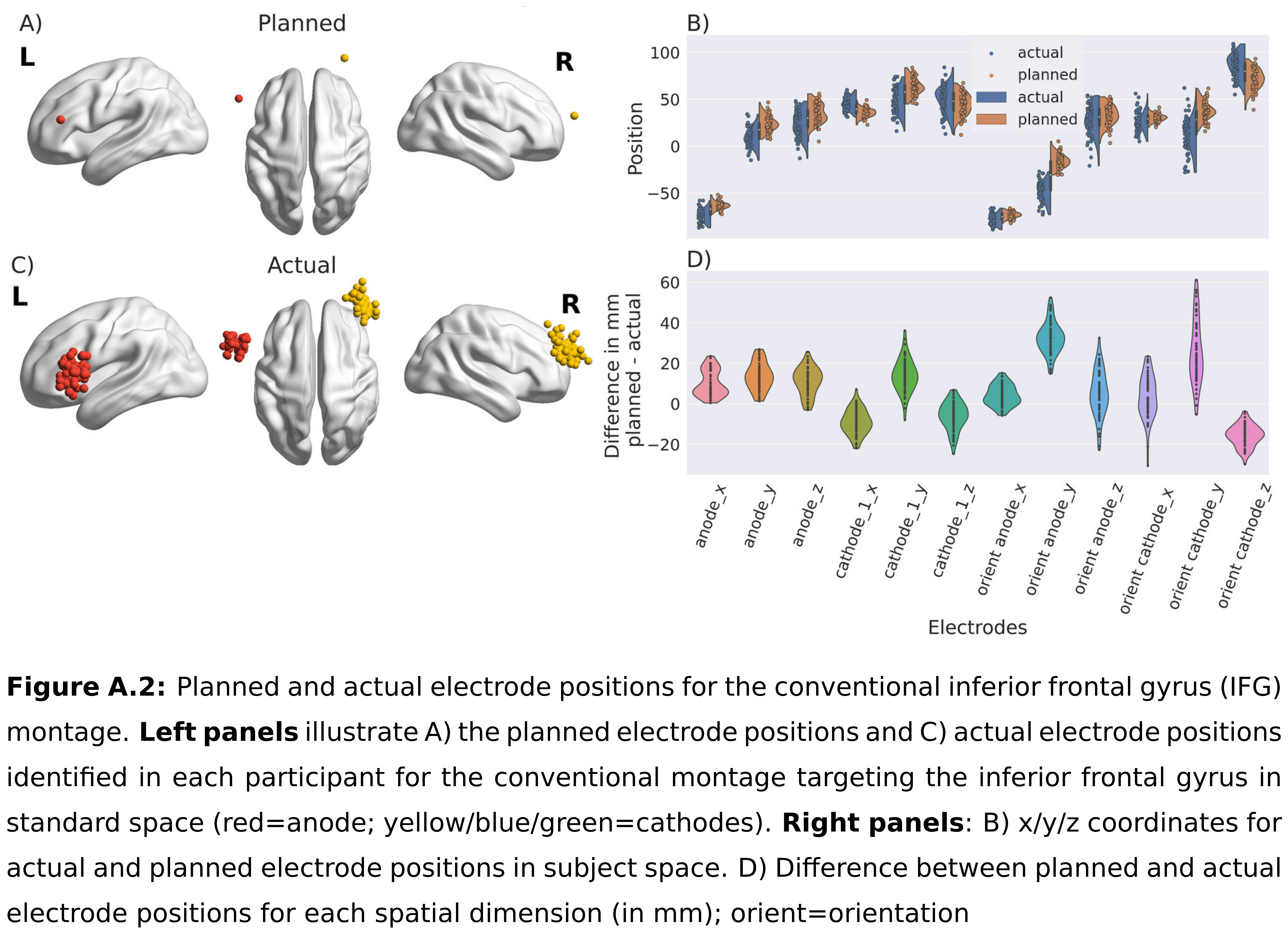

### Appendix Figure 3

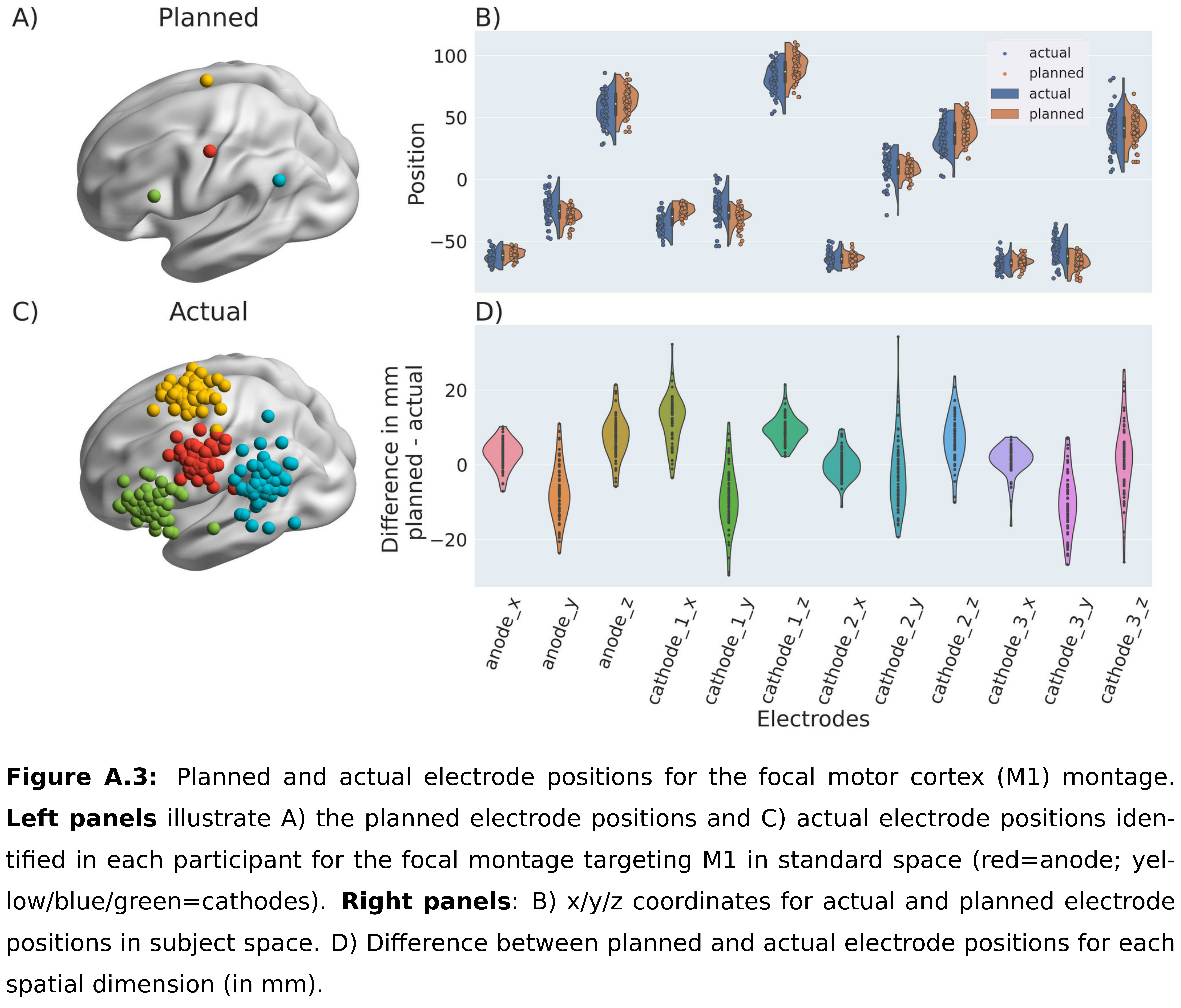

### Appendix Figure 4

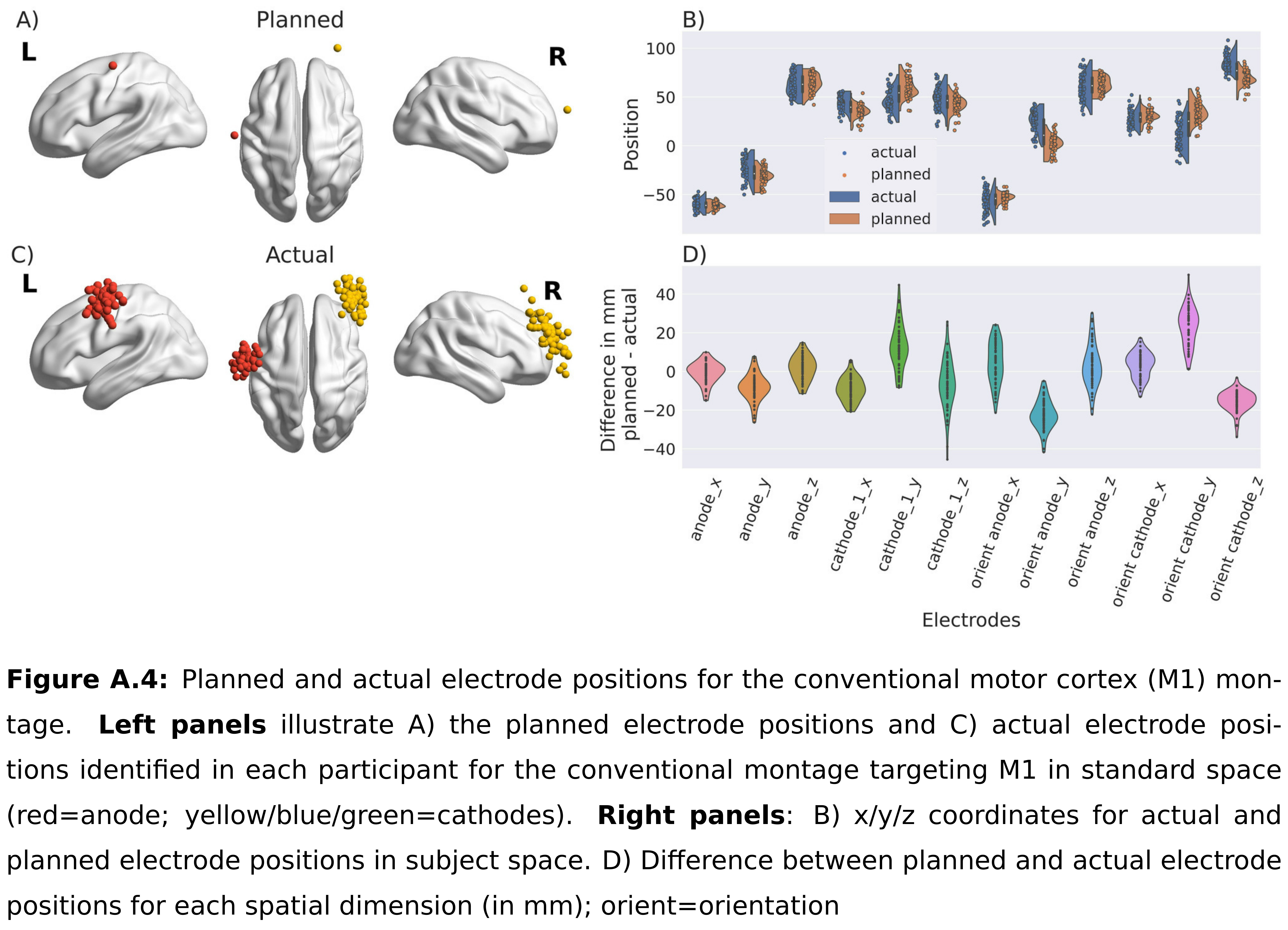

### Appendix Figure 5

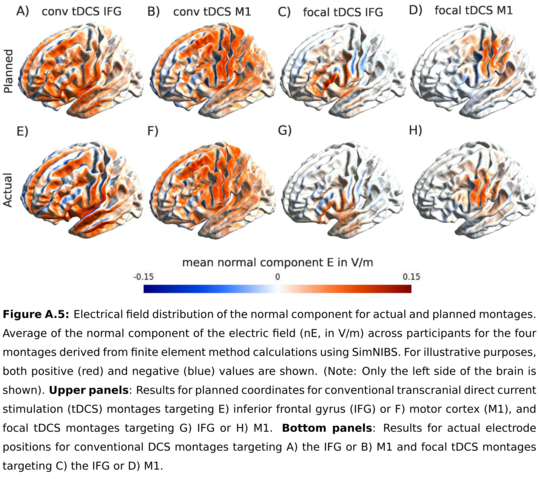

### Appendix Figure 6

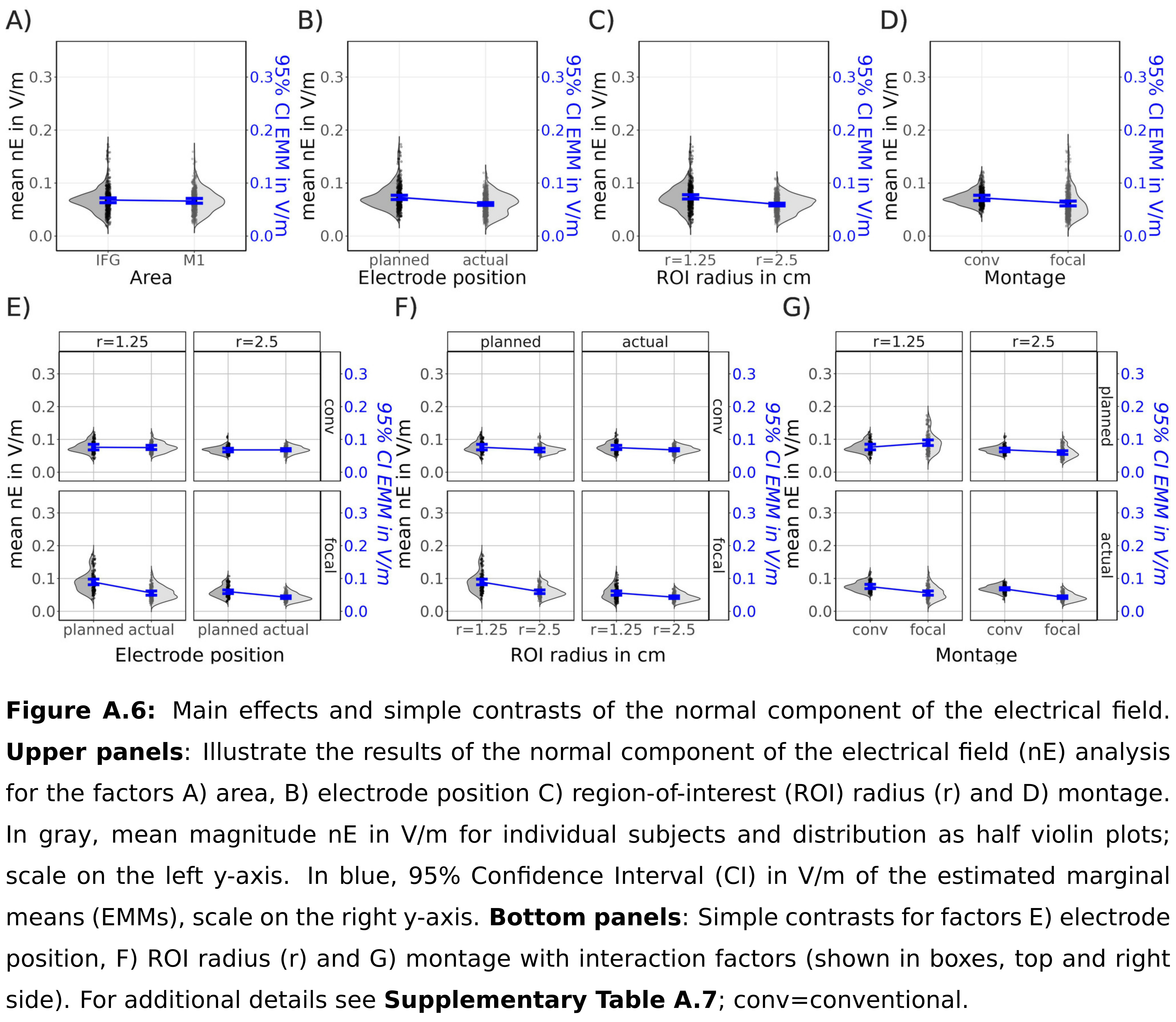

### Appendix Figure 7

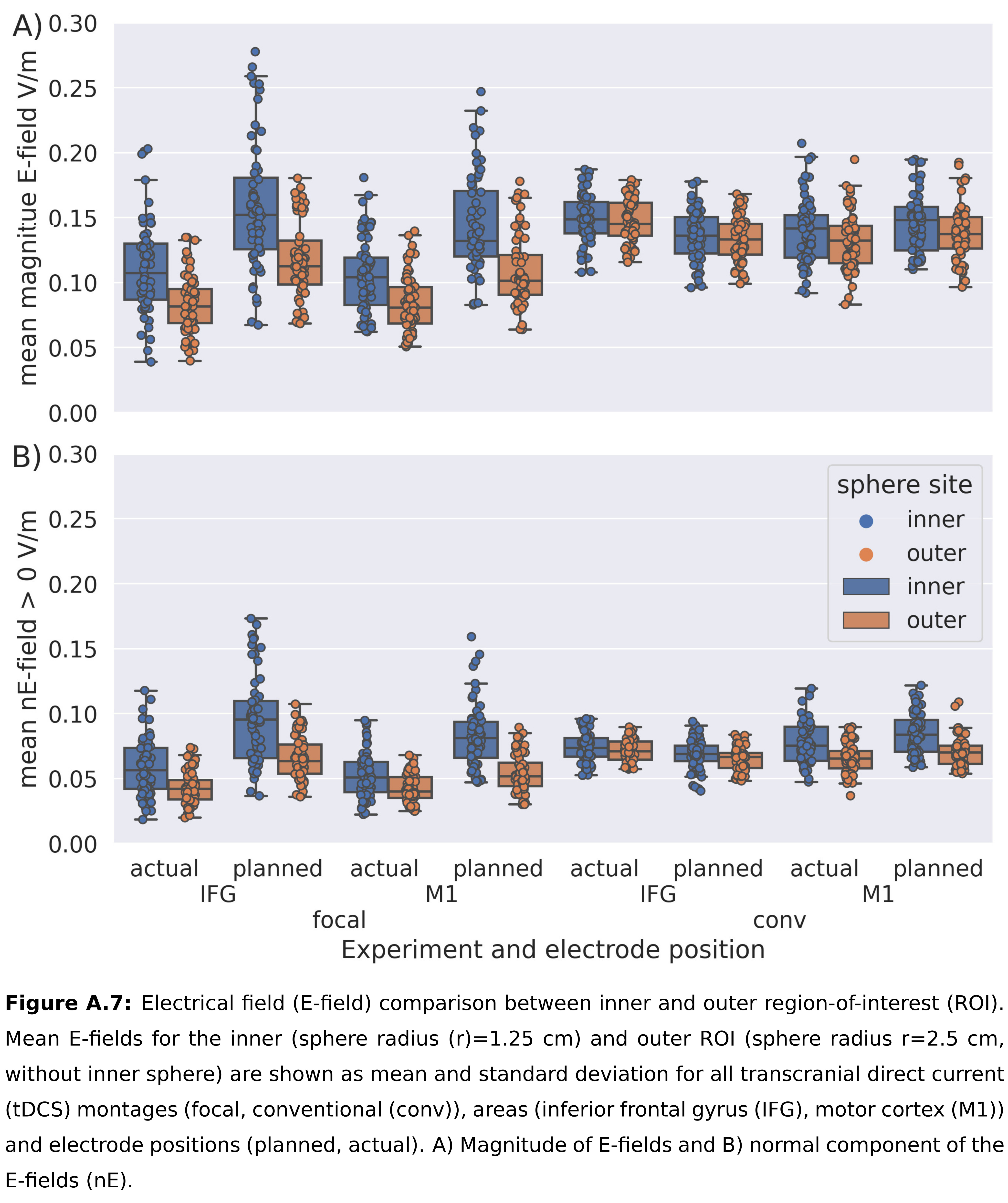
